# What works to support better access to mental health services (from primary care to inpatients) for minority groups to reduce inequalities? A rapid evidence summary

**DOI:** 10.1101/2024.02.28.24303432

**Authors:** Judit Csontos, Deborah Edwards, Elizabeth Gillen, Juliet Hounsome, Meg Kiseleva, Mala Mann, Abubakar Sha’aban, Ruth Lewis, Alison Cooper, Adrian Edwards

## Abstract

It is estimated that one in four people will experience poor mental health throughout their lifetimes. However, ethnic minority groups, refugees and asylum seekers experience more barriers accessing mental health services and have poorer mental health outcomes than those from non-ethnic minority groups. Evidence suggests that interventions that improve access and engagement with mental health services may help reduce disparities affecting ethnic minority groups, refugees and asylum seekers. Thus, the aim of this rapid evidence summary was to explore the literature on what works to support better access to mental health services for ethnic minority groups, refugees and asylum seekers to reduce inequalities. The review included interventions that were developed or assessed to improve equity in access, engagement, utilisation, or provision of mental health services.

**Research Implications and Evidence Gaps:** There is limited review evidence regarding the effectiveness of interventions to improve access to mental healthcare across ethnic minority groups. Review evidence regarding interventions to support refugees and asylum seekers access to primary healthcare or specialised clinics (for example pregnancy and postpartum) is available, but the findings related to mental health care cannot be extracted.

### EXECUTIVE SUMMARY

#### What is a Rapid Evidence Summary?

Our Rapid Evidence Summaries (RES) are designed to provide a rapid response product. They are based on a limited search of key resources and the assessment of abstracts. Priority is given to studies representing robust evidence synthesis. No quality appraisal or evidence synthesis are conducted, and the summary should be interpreted with caution.

This report is linked to a subsequent focused rapid review, to be published in Summer 2024, on the effectiveness of interventions to enhance equitable or overall access to mental health services by ethnic minority groups.

#### Who is this Rapid Evidence Summary for?

Health and Social Services Group - Mental Health & Vulnerable Groups (Policy).

#### Background / Aim of Rapid Evidence Summary

It is estimated that one in four people will experience poor mental health^1^ throughout their lifetimes. However, ethnic minority groups^1^, refugees and asylum seekers^1^ experience more barriers accessing mental health services and have poorer mental health outcomes than those from non-ethnic minority groups. Evidence suggests that interventions that improve access and engagement with mental health services may help reduce disparities affecting ethnic minority groups, refugees and asylum seekers. Thus, the aim of this rapid evidence summary was to explore the literature on what works to support better access to mental health services for ethnic minority groups, refugees and asylum seekers to reduce inequalities. The review included interventions that were developed or assessed to improve equity^1^ in access, engagement, utilisation, or provision of mental health services.

#### Results

##### Recency of the evidence base

▪ The included literature was published between 2006 and 2023.

##### Extent of the evidence base

▪ Bibliographic database searches identified systematic reviews^1^ (n=19), scoping reviews^1^ (n=7), a mapping review^1^ (n=1), narrative reviews^1^ (n=3), overviews of reviews^1^ (n=2), a scoping review of reviews (n=1) and systematic review protocols (n=4).
▪ The stakeholders identified organisational reports (n=4), a rapid review^1^ (n=1), a scoping review (n=1), and a systematic review (n=1).

##### Key findings

▪ The identified literature was summarised under the categories of: Access and pathways to mental health care (separately summarised for ethnic minority groups; Gypsy, Roma and Traveller communities; and refugees and asylum seekers); Mental health promotion, prevention, and treatment of mental health conditions including cultural adaptations to psychological interventions; Help-seeking behaviour; Treatment engagement; and Treatment initiation, participation or continuation.
▪ There is a **wealth of review evidence**, from across the UK and internationally, on the following topics: **barriers and facilitators to accessing mental health care** for ethnic minority groups, refugees and asylum seekers; **mental health promotion, prevention, and treatment** of mental health conditions (including treatment initiation, participation or continuation) for ethnic minority groups, refugees and asylum seekers; and **cultural adaptations** of psychological interventions whether it is for ethnic minority groups living in Western countries or majority populations living in non-Western countries.
▪ While evidence is available on improving mental health care access and experience of ethnic minority groups, refugees and asylum seekers, these often do not have an evaluative component and report their findings as suggestions / recommendations for interventions. Most common **recommendations to improve mental health care equity comprised** of: language and cultural adaptations; education of healthcare professionals; employing ethnically diverse staff; better information provision; collaborative working between different sectors; facilitating referral routes and improving pathways; specialist services for minority groups and outreach services; patient education and skill development; involvement of communities.
▪ Culturally adapted interventions (either mental health promotion, prevention or treatment) may lead to positive outcomes, including improved symptom severity, behaviours, self-efficacy, and wellness, although there is a lack of consensus in what components of adaptations work for which cultural groups. **It is unclear whether cultural adaptations truly result in improved outcomes compared to non-adapted interventions,** and there is a need for high quality well-designed randomised controlled trials.

#### Research Implications and Evidence Gaps

▪ There is limited review evidence regarding the effectiveness of interventions to improve access to mental healthcare across ethnic minority groups.
▪ Review evidence regarding interventions to support refugees’ and asylum seekers’ access to primary healthcare or specialised clinics (for example pregnancy and postpartum) is available, but the findings related to mental health care cannot be extracted.

**Disclaimer:** The views expressed in this publication are those of the authors, not necessarily Health and Care Research Wales. The Health and Care Research Wales Evidence Centre and authors of this work declare that they have no conflict of interest.

## 1. CONTEXT / BACKGROUND

It is estimated that one in four people will experience poor mental health^2^ throughout their lifetimes (Centre for Mental Health 2020). However, international research has documented that ethnic minority groups^2^ experience more barriers to access to mental health services than non-ethnic minorities (Lowther-Payne et al. 2023). At the same time, these groups are at higher risk of mental health conditions^2^ and this risk is often associated with being disproportionally impacted by detrimental social factors, such as racism and poverty (Bignall et al. 2019). Additionally, refugees and asylum seekers^2^ experience higher rates of post-traumatic stress disorder (PTSD) and other mental health disorders linked to dangers associated with migration, poverty, and poor access to health care (Hynie et al. 2023, Iqbal et al. 2022).

Wales is home for diverse ethnic minority groups, including approximately 89,000 people who identify as Asian, Asian British or Asian Welsh), 28,000 Black, Black British, Black Welsh, Caribbean or African individuals, 49,000 mixed or multiple ethnic people, 26,000 members of other ethnic groups (ONS 2022), and 3,630 Gypsy and Irish Traveller residents (ONS 2023). Moreover, around 3,000 asylum seekers are supported in Wales (UK Government 2023a). Together for Mental Health, the Welsh Government’s mental health strategy has focused on ensuring equality since 2012 (Welsh Government 2012). However, the equality impact assessment of this strategy found that since the publication of Together for mental health, stigma and discrimination was still more prevalent for people with protected characteristics, including ethnic minority groups (Welsh Government 2014). In addition, the COVID-19 pandemic highlighted systemic issues, and disproportionately affected ethnic minority groups, prompting the Welsh Government to focus more on reducing health inequalities in their Together for Mental Health updated delivery plan (Welsh Government 2020). A recently published research review of the Together for Mental Health Delivery Plan acknowledged that ethnic minority groups in Wales had poorer mental health outcomes than the wider public while identifying potential barriers to access and service provision (Lock et al. 2023). Improvement in the cultural competency of mental health services and providers in Wales was identified as necessary to help the engagement of ethnic minority groups (Lock et al. 2023). Additionally, issues with multi-lingual service provision in common international languages was also mentioned in the report, which could negatively impact on ethnic minority groups’ help seeking who can discuss their condition better in a language other than English (Lock et al. 2023).

Evidence from the wider international literature also suggests that disparities^2^ in ethnic minority groups’ engagement with mental health services exist along the entire care pathway, with them being less likely to initiate mental health care and fill prescriptions and more likely to end treatment early (Aggarwal et al. 2016, Interian et al. 2013). These disparities are linked to various individual, organisational, and systemic barriers (Aggarwal et al. 2016). Individual-level barriers may include insufficient information to make treatment decisions, communication difficulties and linguistic issues, lack of trust in service providers, psychological distress, fear of stigma, and cultural beliefs resulting in feeling shame about seeking mental health support. Organisational-level barriers refer to unequal access to services and lack of cultural competence in service providers; and the systemic level includes poor funding of mental health services and inaccessibility of information about available services as well as broader issues such as lack of access to transportation or childcare necessary to attend mental health services (Aggarwal et al. 2016). Additionally, barriers to access to and underutilisation of health services, including mental health care, has also been noted among refugees and asylum seekers (Hynie et al. 2023, Iqbal et al. 2022).

To improve ethnic minority groups’ access to mental health services in Wales, a Mental Health Ethnic Minorities Task and Finish Group with the Wales Alliance for Mental Health was set up as part of the Anti-Racist Wales Action Plan (Welsh Government 2022). Moreover, suggestions were made that the new mental health strategy should be developed with the involvement of community organisations, third sector and NHS to make sure that the needs and experiences of ethnic minority groups are considered (Welsh Government 2022). To support these plans, it is also crucial to know what evidence is already available on interventions that could support ethnic minority groups’ access to mental health services. It is thought that effective interventions to improve access and engagement with services may reduce disparities (Interian et al. 2013). Gask et al. (2012) described two conventional approaches to improving access to psychosocial therapies for common mental health problems in underserved groups. Firstly, the ‘default’ position of applying interventions developed for the general population e.g. collaborative care and secondly, modification of existing interventions to make them more acceptable to underserved populations such as developing culturally sensitive psychological therapies. This Rapid Evidence Summary will summarise the available evidence for these two approaches for ethnic minority groups.

## 2. RESEARCH QUESTION(S)

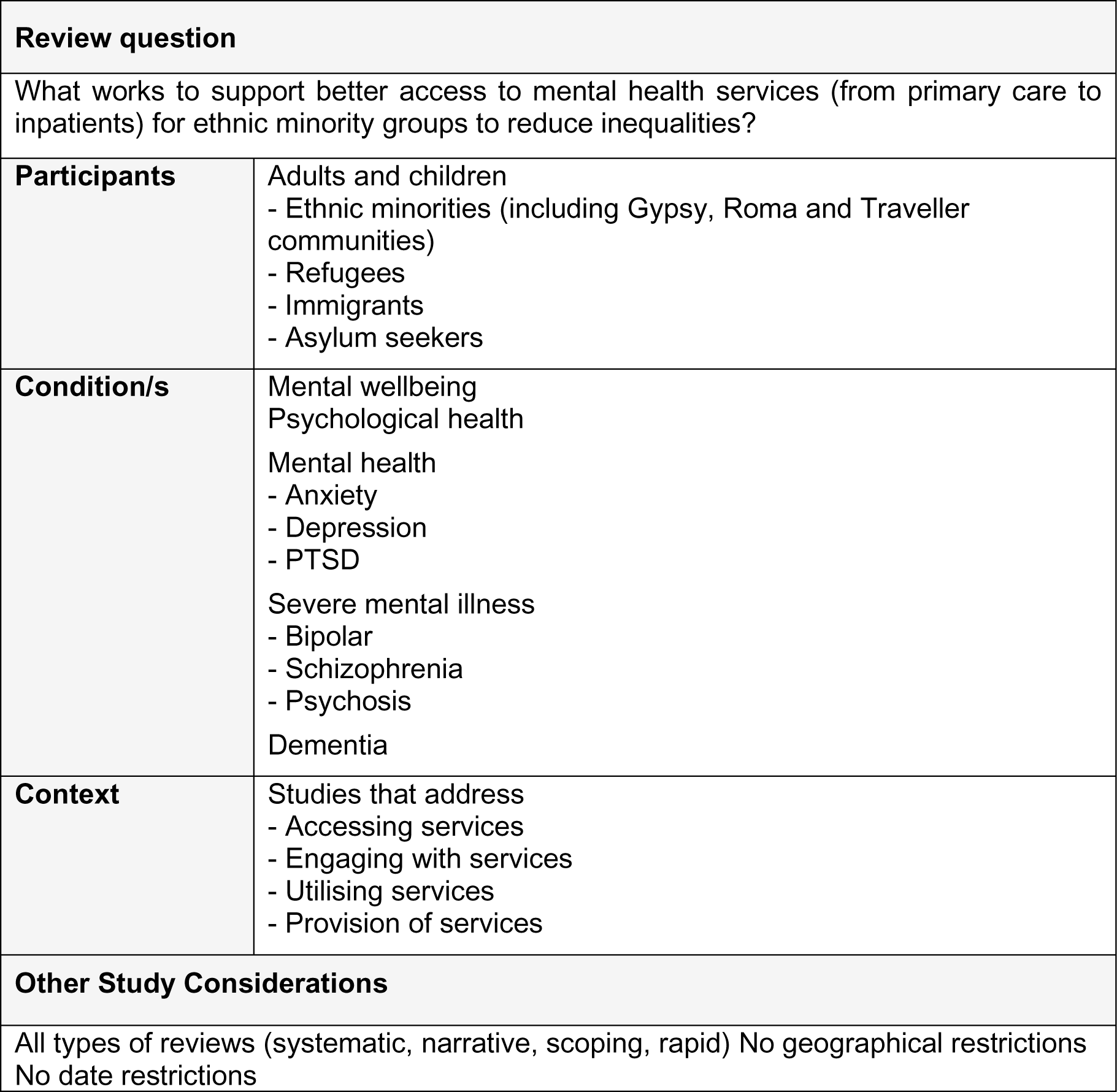

### 1. 3. SUMMARY OF THE EVIDENCE BASE

Secondary research evidence, such as systematic^3^, scoping^3^, and narrative^3^ reviews and overviews of reviews^3^, were identified via searches of various bibliographic databases and the stakeholders. The total number of reports identified is summarised in Table 1 and a summary of each report is provided in Tables 2 to 4 (Section 7). Further detail on the type of evidence identified, their focus and key findings are presented below.

While ethnic minority groups, refugees, asylum seekers, mental health and mental health conditions are defined in the glossary for this rapid evidence summary, included reviews referred to these concepts in different ways. Below the evidence is summarised by using the terms the original authors of the included reviews adopted.

### 3.1. Type and amount of evidence available

**Nineteen** systematic reviews (Aggarwal et al. 2016, Anderson et al. 2015, Arundell et al. 2021, Bhui et al. 2015, Castellanos et al. 2019, Degnan et al. 2018, Ellis et al. 2022, Giebel et al. 2015, Hernandez Robles et al. 2018, Interian et al. 2013, Iqbal et al. 2022, Lee-Tauler et al. 2018, Li et al. 2023, McFadden et al. 2018, Moffat et al. 2009, Moore 2018, Sass et al. 2009, Soltan et al. 2022, Taylor et al. 2023), **seven** scoping reviews (Apers et al. 2023, Handtke et al. 2019, Hynie et al. 2023, Place et al. 2021, Ratnayake et al. 2022, Rogers et al. 2020, Thomson et al. 2015), **one** mapping review^3^ (Arundell et al. 2020), **three** narrative review (Griner and Smith et al. 2006, Kalibatseva and Leong 2014, Mukadam et al. 2013), **two** overviews of reviews (Rathod et al. 2018, Uphoff et al. 2020), **one** scoping review of reviews (Chakanyuka et al. 2022) and **four** protocols (Aslam et al. 2018, Stockwell and Roche 2022, Wang and Kim 2019, Wood et al 2023) were identified across the literature searches of bibliographic databases. The detail from each of these reviews is reported in Table 2.

**Four** organisational reports (Roberts 2023, Traumatic Stress Wales 2023, VCSE Health and Wellbeing Alliance 2022, Welsh Government 2023), **one** rapid review (Kunorubwe et al. 2023), **one** scoping review (Pollard and Howard 2021), and **one** systematic review (Satinsky et al. 2019) were identified by the stakeholder group. The detail from each of these organisational reports and reviews are reported across Tables 3 to 4.

#### 3.1.1 Summary of existing reviews identified via bibliographic databases

This section describes the included reviews and their research aims. Key findings of the included review evidence are summarised in Section 3.2.

The systematic reviews focused on:

- Assessing the relationship between clinician communication and the engagement of racial/ethnic minorities in mental health services across the UK, USA, Australia, Spain and Europe (Aggarwal et al. 2016).
- Assessing the effects of community coalition-driven interventions in improving health status or reducing health disparities among racial and ethnic minority populations of all ages (infant to elderly populations), across the USA, Australia, UK and the Netherlands (Anderson et al. 2015).
- Assessing the efficacy of adapted psychological interventions for people from ethnic minority groups. The specific questions asked were what types of adaptations have been implemented, are the types of culturally adapted interventions differentially effective and what effects of culturally adapted interventions are found across different ethnic minority groups (Arundell et al. 2021).
- Interventions designed to improve therapeutic communications for black and minority ethnic people (young people, adults and the elderly) using psychiatric services across the UK, USA, Canada and Germany (Bhui et al. 2015).
- The effectiveness of mindfulness-based interventions and their cultural adaptations for Hispanic populations from USA, Spain, Chile and Colombia (Castellanos et al. 2019).
- Cultural adaptations of psychosocial interventions for schizophrenia, their effectiveness, specific outcomes and how they were adapted to non-Western populations across 13 different countries (which included ethnic minorities) (Degnan et al. 2018).
- The acceptability and effectiveness of culturally adapted digital mental health interventions for ethnic / racial minorities across the world (Ellis et al. 2022).
- Barriers and facilitators of South Asian older adults’ access to dementia care services in the UK, USA and Canada (Giebel et al. 2015).
- The effectiveness and characteristics of culturally adapted substance use interventions for Latino adolescents in the USA (Hernandez Robles et al. 2018).
- Interventions to improve mental health treatment engagement of underserved racial-ethnic adult groups in the USA (Interian et al. 2013).
- Primary health care interventions to improve the quality of health care (including mental health care) provided to refugees and asylum seekers across OECD countries (Iqbal et al. 2022).
- Interventions to improve the initiation of mental health care among racial-ethnic minority groups in the USA (Lee-Tauler et al. 2018).
- The effectiveness of culturally adapted treatments for common mental disorders in people of Chinese descent, including those living in mainland China and in other countries as ethnic minorities (Li et al. 2023).
- Addressing the question of how Gypsy, Roma and Traveller populations access healthcare and what are the best ways to enhance their engagement with health services (including mental health services), in various European countries and Canada, with about half of the evidence conducted in or including the UK (McFadden et al. 2018).
- Improving pathways to mental health care for adults and young people from black and ethnic minority groups in the UK (Moffat et al. 2009).
- Interventions to improve engagement with mental health services (initiation, participation, completion and service utilisation) among adolescents and young adults from underserved minority groups in the USA (Moore 2018).
- Initiatives to enhance pathways to mental health care for adults from black and ethnic minority populations across, UK, Australia and USA (Sass et al. 2009).
- The effectiveness and acceptability of community-based interventions based on randomised controlled trials (RCTs) only in comparison with controls (no treatment, waiting list, alternative treatment) for preventing and treating mental health problems (major depression, anxiety, PTSD, psychological distress) and improving mental health in refugee children and adolescents in high-income countries (Soltan et al 2022).
- Determining the efficacy of culturally adapted psychosocial interventions in comparison to generic treatment for young people with refugee or asylum-seeker status in the UK (Taylor et al. 2023)

The scoping reviews focused on:

- Health promotion, prevention, and non-medical treatment interventions targeting the mental health and mental wellbeing of migrants and ethnic minority groups in Europe (Apers et al. 2023).
- To identify and evaluate intervention components and strategies that create culturally competent healthcare (including mental health care) for culturally and linguistically diverse people and to develop a model of culturally competent healthcare provision (Handtke et al. 2019).
- The accessibility (affordability, availability/accommodation, appropriateness and acceptability) of virtual mental healthcare interventions and assessments for refugee and immigrant groups of all ages globally across multiple countries and settings (Hynie et al. 2023).
- Mapping the interventions to increase migrants’ care-seeking behaviour in high-income countries for stigmatised conditions with a primary focus on mental health (Place et al. 2021).
- Mapping the types and characteristics of approaches and interventions that non-medical local immigrant settlement organisations undertake to support access to primary healthcare and mental health services for immigrants over 16 years of age in high-income countries (Canada, USA, Australia) (Ratnayake et al. 2022).
- Models of care in pregnancy and the first 12 months postpartum in migrant and refugee women living in high income countries, including but not limited to mental health care (Rogers et al. 2020).
- Barriers to immigrant populations utilisation of mental health services in Canada and analysis of policy and practice recommendations (Thomson et al. 2015).

The mapping review explored interventions that aim to reduce mental health inequalities, their cost-effectiveness and the barriers and facilitators affecting them (Arundell et al. 2020). This mapping review included populations from multiple protected characteristics, including race, religion and belief (Arundell et al. 2020).

The narrative reviews focused on:

- Determining the effectiveness of culturally adapted mental health interventions by using meta-analysis, although the methods of literature reviewing did not resemble a standard systematic review. The countries where the research was conducted were not reported (Griner and Smith 2006).
- Critically examining the literature on culturally sensitive treatments for depression. The countries where the research was conducted were not reported (Kalibatseva and Leong 2014).
- Reasons for underutilisation of dementia services by minority ethnic groups and the interventions that have been developed to address access issues. The countries where the research was conducted were not reported (Mukadam et al. 2013).

The overviews of reviews focused on:

- Summarising systematic and literature reviews with meta-analyses which aimed to determine the effectiveness of cultural adaptations of mental health interventions. The countries where the research was conducted were not reported (Rathod et al. 2018).
- Mapping the characteristics and methodological quality of existing systematic reviews and registered systematic review protocols on the promotion of mental health and prevention and treatment of common mental disorders among refugees, asylum seekers, and internally displaced persons. The countries where the research was conducted were not reported (Uphoff et al. 2020).

The objective of the scoping review of reviews was to appraise the existing literature to identify key elements, conceptualisations, and interventions of cultural safety to improve health services and dementia care for Indigenous people from Canada, Australia and New Zealand (Chakanyuka et al. 2022).

The systematic review protocols focused on:

- Barriers and facilitators to the uptake of psychosocial intervention that are delivered by lay therapists to improve asylum seekers’ and migrants’ mental health and wellbeing (Aslam et al. 2018).
- Culturally sensitive interventions for black men to improve access to mental health care and/or mental health outcomes in the community (Stockwell and Roche 2022).
- Community-based interventions used to address Asian Americans’ mental health challenges (Wang and Kim 2019).
- Community-level interventions for people from minority ethnic groups to improve their access to primary care when they experience a first-episode psychosis (Wood et al. 2023).

#### 3.1.2 Summary of organisational reports and existing reviews identified via stakeholder group

These explored the following:

- The rapid review aimed to provide up-to-date evidence on accessibility, appropriateness, acceptability and outcomes of psychological interventions for people from Black, Asian and minority ethnic communities and to provide evidence of both good practice and barriers to good practice for all ages groups in the UK (Kunorubwe et al. 2023).
- The scoping review explored barriers and enablers of mental health care for asylum seekers and refugees residing in the UK (Pollard and Howard 2021).
- The systematic review investigated the barriers to mental health care utilisation and access among refugees and asylum seekers in Europe (Satinsky et al. 2019).
- The scope of the evidence on the management of PTSD for refugees and asylum seekers, with a focus on psychological interventions (Roberts 2023).
- A scoping exercise identified good practice in Wales and across the UK with regard to access to psychological therapies and mental health services for Asylum Seekers, Refugees and Migrants in the UK (Traumatic Stress Wales 2023).
- A qualitative primary research report regarding the barriers and opportunities for improving access to mental health support for refugees and people seeking asylum in England (VCSE Health and Wellbeing Alliance 2022).
- A qualitative primary research report into “Good Access” in Community Pharmacy, NHS Dentistry and Allied Health Professional Services which included barriers and facilitators to access (Welsh Government 2023).

### 3.2. Key Findings

The identified literature was categorised based on the aspect of mental health care access and provision it investigated. The categories are: Access and pathways to mental health care; Mental health promotion and prevention, and treatment of common mental disorders; Cultural adaptations to psychological interventions; Help seeking behaviour; Treatment engagement; and Treatment initiation, participation or continuation. The key findings of the identified literature are briefly summarised below under each category. The findings relating to Access and pathways to mental health care for minority groups are reported separately for ethnic minority groups; Gypsy, Roma and travelling communities; and migrants, refugees and asylum seekers. Key findings from the literature identified by the stakeholders were narratively summarised if the reports focused on interventions aiming to improve access to services or treatment modalities.

#### Access and pathways to mental health care for ethnic minority groups

- Barriers to **South Asian older adults’** access to dementia care include insufficient knowledge about dementia, mental illness, and local services, stigma, culturally preferred coping strategies, and linguistic and cultural difficulties in communication. **Suggestions were provided that could help improve access** for South Asian older adults and these included better information provision, culturally tailored services, employment of healthcare staff with similar cultural background, and education of healthcare professionals about dementia and local services (Giebel et al. 2015).
- The key components of **effective pathway interventions to mental health care** for **black and ethnic minority groups** included specialist services for ethnic minority groups, collaboration between sectors, facilitating referral routes between services, outreach and facilitating access into care, and supporting access to rehabilitation and moving out of care (Moffat et al. 2009).
- Sass et al. (2009) reported evidence of interventions with **black and ethnic minority populations** that led to three types of pathway modification (accelerated transit through care pathways, removal of adverse pathways, the addition of beneficial pathways). It was found that ethnic matching promoted desired pathways in some ethnic groups, that managed care improved equity, that a pre-treatment service improved access to detoxification, and that an education leaflet increased recovery.

#### Access and pathways to mental health care for people from Gypsy, Roma and Traveller communities

- The barriers to health services (including mental health services) reported by people from **Gypsy, Roma and Traveller communities** included the organisation of health systems, discrimination, culture and language, health literacy, service-user attributes, and economic barriers. Strategies thought to improve engagement included specialist roles, outreach services, dedicated services, raising health awareness, handheld records, training for staff, and collaborative working (McFadden et al. 2018).

#### Access and pathways to mental health care for refugees and asylum seekers

- The scoping review by Hynie et al. (2023) described factors affecting accessibility of **virtual mental health interventions** for **refugee and immigrant groups** which were individual (e.g., literacy), program related (e.g., computer needed), and/or contextual/social (e.g., housing characteristics, internet bandwidth). Need for financial and technical support were identified as potential barriers to refugee and immigrant groups’ access.
- The interventions provided to **refugees and asylum seekers** within the broad primary care setting to improve the quality of health care, spanning mental health (n=11) and general wellbeing (n=44) could be organized into four categories, for example those that focused on developing the skills of individual refugees/asylum seekers and their families (**including promoting access** and improving engagement with and adherence to health regimes); skills of primary health care workers; system and/or service integration models and structures; and lastly, interventions enhancing communication services (Iqbal et al. 2022).
- Findings from the scoping review by Ratnayake et al. (2022) showed that local non-medical **immigrant** settlement organisations had established approaches and interventions to support immigrants **access to primary healthcare services**, with most studies showing that mental health support was an important component. These included connecting to healthcare services and / or collaborating with health sector institutions; providing health promotion programs; undertaking community capacity-building and policy advocacy activities and providing on the ground assistance.
- The scoping review by Rogers et al. (2020) identified a range of **interventions that improved access to maternity and postpartum care for migrant and refugee women**, including bilingual/bicultural workers, group antenatal care, and specialised clinics for care in pregnancy or first 12 months postpartum. **Four** of the 17 included studies had a mental health focus but these did not report separate findings.
- Barriers to **immigrant populations’** utilisation of mental health services in Canada included the uptake of health information, the process of immigrant settlement, and service availability. **Recommendations to improve access** included linguistically and culturally sensitive services, provision of translators and training for mental health workers in cultural and language competencies, practitioners considering patients’ circumstances and pre-immigration experiences, diverse and socially inclusive workforce, and involvement of the community among other suggestions (Thomson et al. 2015).

#### Mental health promotion, prevention and treatment of mental health conditions including cultural adaptations to psychological interventions

- The review by Anderson et al. (2015) found one high-quality study which was a cluster-randomised trial that implemented **depression care quality improvement** in a network of mental health and health and social care systems (primary care, substance abuse, social services, and homeless services) in the ethnically diverse South Los Angeles and Hollywood metropolitan area, USA. That study found added benefit of a community coalition-driven intervention^4^ for improved mental health among **African Americans.**
- The scoping review by Apers et al. (2023) identified three main **successful mechanisms for intervention development and implementation** with regard to the **prevention and promotion** of mental health and wellbeing, secondary prevention and tertiary prevention for **migrants and ethnic minority** groups: a sound theory-base, systematic adaption to make interventions culturally sensitive and participatory approaches.
- The mapping review reported that 80% of their identified studies aimed at improving inequalities in people from lower socioeconomic groups, diverse age groups or **ethnic minority and indigenous/aboriginal communities**. The interventions identified included **psychological support, delivering education and training, engaging the community and other culturally adapted interventions** (Arundell et al. 2020).
- **Cultural adaptations** made to psychological interventions for **ethnic minority groups** with mental health problems appear to be efficacious relative to non-adapted or waitlist/no intervention comparators. Adaptions included adapting the form used to provide interventions, the time or length of the intervention, the location of treatment provision, and the method of access (Arundell et al. 2021).
- The review by Bhui et al. (2015) reports on beneficial interventions to improve **therapeutic communications** for **black and minority ethnic people** receiving specialist psychiatric care. The interventions with evidence of benefit were culturally adapted psychotherapies (cognitive–behavioural therapy and family therapies); ethnographic and motivational interviewing; communications skills training; community-based stepped care and case finding by including social venues in the care pathway; role induction and education for patients and telepsychiatry that included ethnic matching.
- Findings from a meta-analysis indicate that **cultural adaptation of mindfulness-based interventions** for **Hispanic populations** might improve depressive symptoms and stress and could help with managing a chronic illness (Castellanos et al. 2019). A moderate to large effect could also be detected on psychiatric distress. However, the authors conclude that more high quality research studies were needed.
- The scoping review of reviews found little evidence that reported on interventions that aimed to improve **cultural safety in health and dementia care** for **Indigenous people** (Chakanyuka et al. 2022). There was also a lack of research focusing on implementation and evaluation of cultural safety intervention, and using the concept of sex and gender (Chakanyuka et al. 2022).
- Psychosocial interventions for schizophrenia can be culturally adapted based on language, concepts and illness models, family, communication, content, cultural norms and practices, context and delivery, therapeutic alliance, and treatment goals. Findings from the meta-analysis of **culturally adapted psychosocial interventions** for **people from non-Western countries** imply reduced symptom severity, although change was proportionate to level of adaptation. Nevertheless, there is a lack of well-designed high quality studies comparing adapted interventions to non-adapted (Degnan et al. 2018).
- **Culturally adapted digital mental health interventions** for **ethnic / racial minorities** have been found effective and acceptable in outcomes such as, mental health symptomatology, behaviours, self-efficacy, or wellness, although there are a lack of studies focusing on feasibility, active comparison treatments or on Black and Indigenous groups (Ellis et al. 2022).
- A narrative review containing a meta-analysis indicates that **cultural adaptations of mental health interventions to specific populations** are four times more effective than interventions aimed at mixed cultural or ethnic groups. In addition, intervention adapted to patients’ mother tongue were twice as effective as English language treatments (Griner and Smith 2006).
- A scoping review focusing on **culturally competent healthcare** (including mental health care) for **culturally and linguistically diverse people** identified 20 categories of different strategies, which could be organised into four groups: Components of culturally competent healthcare–Individual level; Components of culturally competent healthcare–Organizational level; Strategies to implement culturally competent healthcare; and Strategies to provide access to culturally competent healthcare (Handtke et al. 2019). Both qualitative and quantitative findings indicate positive patient and service utilization outcomes as a result of culturally competent healthcare strategies, although the effect was often small or not statistically significant (Handtke et al. 2019).
- A meta-analysis of **culturally adapted substance use interventions** indicates that the substance use outcomes of **Latino adolescents** in the USA are slightly improved, although the quality of included studies was low (Hernandez Robles et al. 2018).
- **Culturally sensitive treatments** for **people with depression** can contain general or practical adaptations, including translation of written materials or incorporating cultural values (Kalibatseva and Leong 2014).
- A rapid review reports that both **culturally sensitive and culturally adapted psychological interventions** have shown positive results in terms of accessibility, appropriateness, acceptability and outcomes amongst **Black, Asian and minority ethnic communities**. It is of note however, that there is still limited research on the efficacy of such interventions across distinct cultural communities. The literature also supports the utilisation of more community based collaborative approaches to service development to remove access barriers and improve outcomes (Kunorubwe et al. 2023).
- **Culturally adapted interventions** for **Chinese descents with common mental disorder** have been found moderately effective in reducing symptom severity, although no difference was detected between culturally modified and culturally specific interventions. However, the quality of included studies could not be determined due to issues with reporting (Li et al. 2023).
- An overview of reviews focusing on **cultural adaptation of mental health interventions** highlighted that there is a lack of consensus regarding what components of adaptations work for which cultural groups. Additionally, it is unclear whether cultural adaptations truly result in improved outcomes compared to non-adapted interventions (Rathod et al. 2018).
- Roberts (2023) scoped the evidence on the **management of PTSD** for **refugees and asylum seekers**. The author reported on five RCTs, a network meta-analysis (n=23 studies) of psychosocial interventions for refugees and asylum seekers with PTSD (Turrini et al. 2021) and an individual-patient data meta-analysis of residual PTSD symptoms after provision of brief behavioural intervention in low- and middle-income countries (Akhtar et al. 2022). Cognitive behavioural therapy and Eye movement desensitisation and reprocessing were found significantly more effective in reducing PTSD symptoms than waitlist comparison (Turrini et al. 2021). Problem Management Plus^5^ intervention was also observed to be more effective for symptom reduction than enhanced treatment as usual, although residual symptoms were reported for 30% of refugee and asylum seeker participants (Akhtar et al. 2022).
- There was no evidence of an effect of **community-based interventions** when compared with a waiting list for symptoms of PTSD, depression, and psychological distress in **refugee children and adolescents** in high-income countries (Soltan et al. 2022).
- A systematic review investigating **cultural adaptations** to psychosocial interventions for **young people with refugee or asylum-seeker status** found that these type of interventions may have some positive impact on symptom reduction, although results varied. However, the available evidence is insufficient to determine effectiveness of cultural adaptations compared to treatment as usual or to investigate which components of cultural adaptation work better, as study designs were mainly qualitative or observational (Taylor et al. 2023).
- The overview of reviews mapped the evidence-base and found that there were gaps in the literature regarding mental health interventions for refugees, asylum seekers and internally displaced persons. Existing systematic reviews focus more attention on the treatment of PTSD than mental health promotion or prevention, or the treatment of depression or anxiety. Studies of Cognitive Behavioural Therapy, Narrative Exposure Therapy, and integrative and interpersonal therapies were most likely to be included in reviews (Uphoff et al 2020).

#### Help seeking behaviour

- Educational campaigns to increase awareness and reduce stigma of dementia for **minority ethnic groups** have been described in the literature, although outcomes of these interventions have not been reported. However, other studies suggest that information targeted to specific ethnic groups has the potential to increase knowledge about dementia and consequently help-seeking (Mukadam et al. 2013).
- The scoping review by Place et al. (2021) identified three approaches that increased **migrants’** mental health care-seeking behaviour: health communication (n=9); support groups (n = 2); and primary care-based approaches (n=4), including a self-assessment tool for psychosocial risk, and engagement interventions seeking to increase collaboration between primary care and mental health services.

#### Treatment engagement

- The review by Interian et al. (2013) found that collaborative care for depression was efficacious as an engagement improvement intervention for **underserved racial-ethnic groups**.

Interventions that were family based or were culturally adapted for age group or race-ethnicity showed possible efficacy and promising results for improving treatment engagement among **underserved minority adolescents and young adults** (Moore 2018).

#### Treatment initiation, participation or continuation

- The review by Aggarwal et al. (2016) did not find any RCTs of interventions to target specific mechanisms of action for improving patient-clinical communication regarding treatment initiation, participation or continuation with **racial/ethnic minorities**. Although some studies were found that reported on clinician experiences to improve patient-clinician communication.
- Interventions to improve the initiation of mental health care among **racial-ethnic minority groups** included collaborative care (n=10), psychoeducation (n=7), case management (n=5), co-location of mental health services within existing services (n=4), screening and referral (n=2) and a change of Medicare medication reimbursement policy (n=1). An increased uptake of psychotherapy or antidepressant use among members of racial-ethnic minority was observed across interventions involving either collaborative care, co-location of mental health services, and screening and referral interventions (Lee-Tauler et al. 2018).

### 3.3. Areas of uncertainty

- There is a wealth of review and overview of review evidence that explores the barriers and facilitators to accessing mental healthcare for ethnic minority groups, refugees and asylum seekers across the UK and internationally. However, these often do not have an evaluative interventions and report their findings as suggestions / recommendations for interventions.
- There is a wealth of review evidence that covers mental health promotion, prevention and treatment of mental health conditions (including treatment initiation, participation or continuation) for ethnic minority groups, refugees and asylum seekers across the UK and internationally.
- There is a wealth of review and overview of review evidence focusing on cultural adaptations of psychological interventions whether it is for ethnic minority groups living in Western countries or majority populations living in non-Western countries.
- There is limited review evidence regarding the effectiveness of interventions to improve access to mental health care across ethnic minority groups.
- There is wider review evidence regarding interventions to support refugees and asylum seekers access to primary healthcare services and specialised clinics for care in pregnancy and postpartum. However these reviews did not separate out the findings for the studies that focused on mental health.

## 4. NEXT STEPS

The findings of this Rapid Evidence Summary were shared with the stakeholders and used to inform our decision on a substantive focus for a subsequent Rapid Review (RR). It was decided that the RR would focus on interventions to support equitable access to mental health services for ethnic minority groups.

## Data Availability

All data produced in the present study are available upon reasonable request to the authors

## Abbreviations

BAME: Black, Asian and minority ethnic
NHS: National Health Service
OECD: Organisation for Economic Co-operation and Development
ONS: Office for National Statistics
PTSD: Post-traumatic stress disorder
RCT: Randomised Control Trial
VCSE: Voluntary Community and Social Enterprise

## Glossary

<u>Disparity:</u> “Health disparity and health inequality are broad terms that include health inequity and signify more than just difference or variation: they signify a health difference that raises moral or ethical concerns.” (Braveman et al. 2018, p. 11)

<u>Equity:</u> Health equity means that “everyone has a fair and just opportunity to be as healthy as possible. Achieving health equity requires removing obstacles to health such as poverty, discrimination, and their consequences, which include powerlessness and lack of access to good jobs with fair pay; quality education, housing, and health care; and safe environments. For the purposes of measurement, health equity means reducing and ultimately eliminating disparities in health and health determinants that adversely affect excluded or marginalized groups. Health equity is the ethical and human rights principle motivating efforts to eliminate health disparities; health disparities are the metric for assessing progress toward health equity.” (Braveman et al. 2018, p. 11)

<u>Ethnic minorities:</u> The term refers to “all ethnic groups except the white British group. Ethnic minorities include white minorities, such as Gypsy, Roma and Irish Traveller groups.” (UK Government 2023b)

<u>Mental health:</u> Welsh Government defines mental health as a state of mental wellbeing that enables people to cope with the stresses of life, realise their abilities, learn well and work well, and contribute to their community. It is an integral component of health and wellbeing that underpins our individual and collective abilities to make decisions, build relationships and shape the world we live in. Mental health is a basic human right. And it is crucial to personal, community and socio-economic development. People with poor mental health can have a mental health condition but this is not always or necessarily the case.

<u>Mental health condition:</u> Welsh Government defines mental health condition as a broad term covering conditions that affect emotions, thinking and behaviour, and which substantially interfere with our life. Mental health conditions can significantly impact daily living, including our ability to work, care for ourselves and our family, and our ability to relate and interact with others. This is a term used to cover several conditions (e.g. depression, post-traumatic stress disorder, schizophrenia) with different symptoms and impacts for varying lengths of time, for each person. Mental health conditions can range from mild through to severe and enduring illness. People with mental health conditions are more likely to experience lower levels of physical and mental wellbeing, but this is not always or necessarily the case. Some mental health conditions like eating disorders and schizophrenia are associated with a higher risk of mortality.

<u>Refugees and asylum seekers:</u> “Refugees are people forced to flee their own country and seek safety in another country. They are unable to return to their own country because of feared persecution as a result of who they are, what they believe in or say, or because of armed conflict, violence or serious public disorder” (UNHCR 2024b). “An asylum-seeker is someone whose request for sanctuary has yet to be processed” (UNHCR 2024a).

Different review types:

<u>Mapping review:</u> “is a transparent, rigorous, and systematic approach to identifying, describing, and cataloguing evidence and evidence gaps in a broader topic area.” (Campbell et al. 2023, p. 5)

<u>Narrative review:</u> Often referred to as literature review, can be defined as a “generic term for published materials that provide examination of recent or current literature, can cover wide range of subjects at various levels of completeness and comprehensiveness. It may include research findings.” (Grant and Booth 2009, p. 94)

<u>Overview of reviews:</u> “uses explicit and systematic methods to search for and identify multiple systematic reviews on related research questions in the same topic area for the purpose of extracting and analysing their results across important outcomes. Thus, the unit of searching, inclusion and data analysis is the systematic review.” (Pollock et al. 2023)

<u>Rapid review:</u> “a type of knowledge synthesis in which systematic review processes are accelerated and methods are streamlined to complete the review more quickly than is the case for typical systematic reviews.” (Tricco et al. 2017, p. 3)

<u>Scoping review:</u> “a type of evidence synthesis that aims to systematically identify and map the breadth of evidence available on a particular topic, field, concept, or issue, often irrespective of source (i.e., primary research, reviews, non-empirical evidence) within or across particular contexts. Scoping reviews can clarify key concepts/definitions in the literature and identify key characteristics or factors related to a concept, including those related to methodological research.” (Munn et al. 2022, p. 950)

<u>Systematic review:</u> “A systematic review attempts to identify, appraise and synthesize all the empirical evidence that meets pre-specified eligibility criteria to answer a specific research question. Researchers conducting systematic reviews use explicit, systematic methods that are selected with a view aimed at minimizing bias, to produce more reliable findings to inform decision making.” (Cochrane Library 2024)

## 6. RAPID EVIDENCE SUMMARY METHODS

A list of the resources searched during this Rapid Evidence Summary is provided within the Appendix. Relevant research was also identified via the stakeholders. Searches were limited to English-language publications and did not include searches for primary studies. Search hits were screened for relevance by a single reviewer.

Priority was given to robust evidence synthesis using minimum standards (systematic search, study selection, quality assessment, appropriate synthesis). The secondary research identified was not formally quality assessed. The included research may vary considerably in quality and the degree of such variation could be investigated during rapid review work which may follow-on. Citation, recency, evidence type, document status and key findings were tabulated for all relevant secondary research identified in this process.

|  |  |
| --- | --- |
| **Date of Search** | November 2023 |
| **Search Concepts Used** | **Mental health** (synonyms and related terms: psychological health, mental illness, bipolar, schizophrenia, psychosis, depression, anxiety, post-traumatic stress disorder, dementia)<br>AND<br>**Minority groups** (synonyms and related terms: gypsy, roma, traveller, refugee, asylum, ethnic, minority, racial, immigrant)<br>AND<br>**Intervention** (synonyms and related terms: improve)<br>AND<br>**Access** (synonyms and related terms: provision, service, engage) |
| **Search Completed by** | Cardiff Evidence Synthesis Collaborative |

## 7. EVIDENCE

**Table 1:** Summary of review evidence identified. A more detailed summary of included evidence can be found across Table 2 to 4.

| **Evidence type** | **Total identified** |
| --- | --- |
| Systematic reviews | 20 |
| Scoping reviews | 8 |
| Mapping review | 1 |
| Rapid reviews | 1 |
| Narrative reviews | 3 |
| Overview of reviews | 2 |
| Scoping review of reviews | 1 |
| Protocols for reviews that are underway | 4 |
| Stakeholder organisational reports | 4 |

**Table 2:**
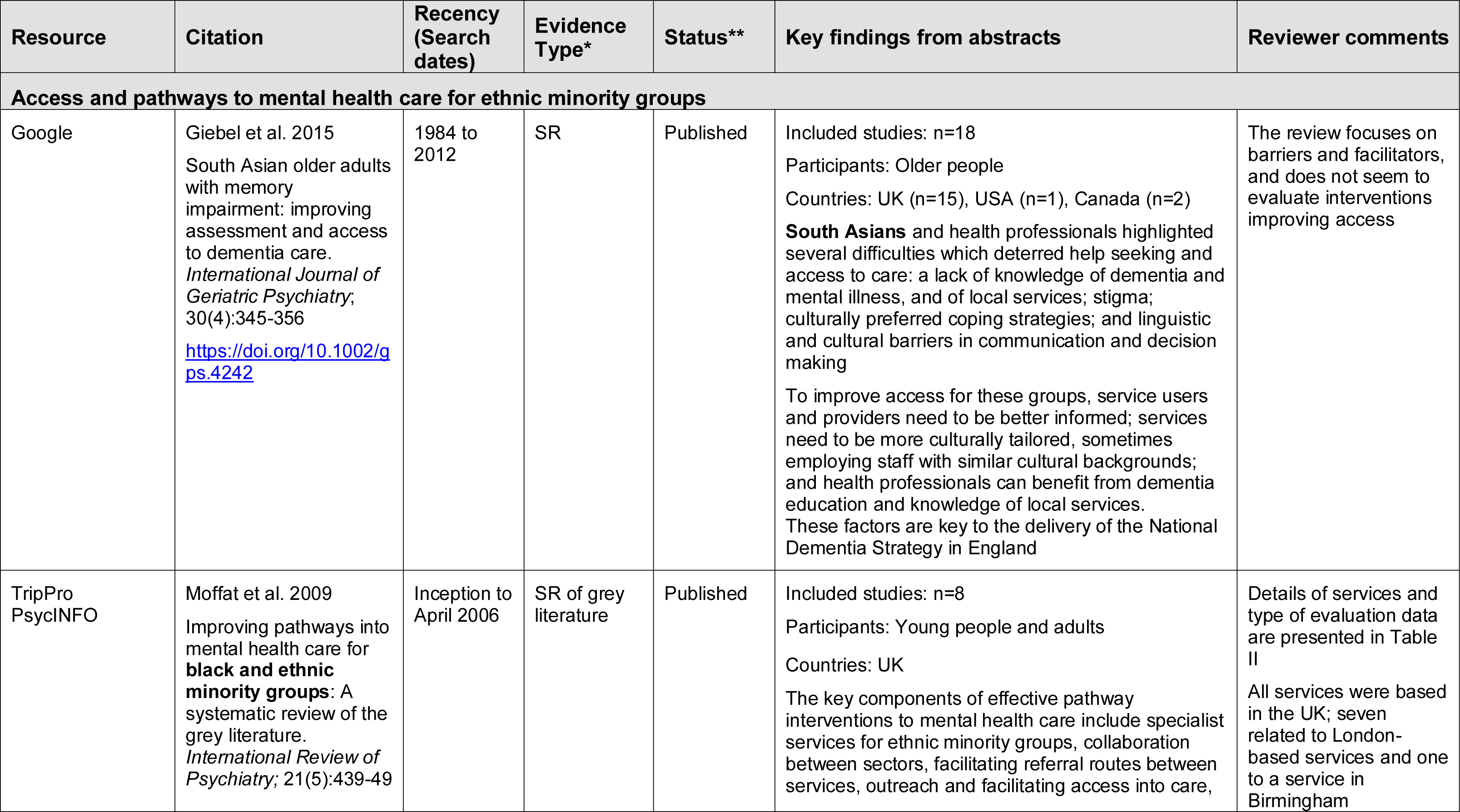
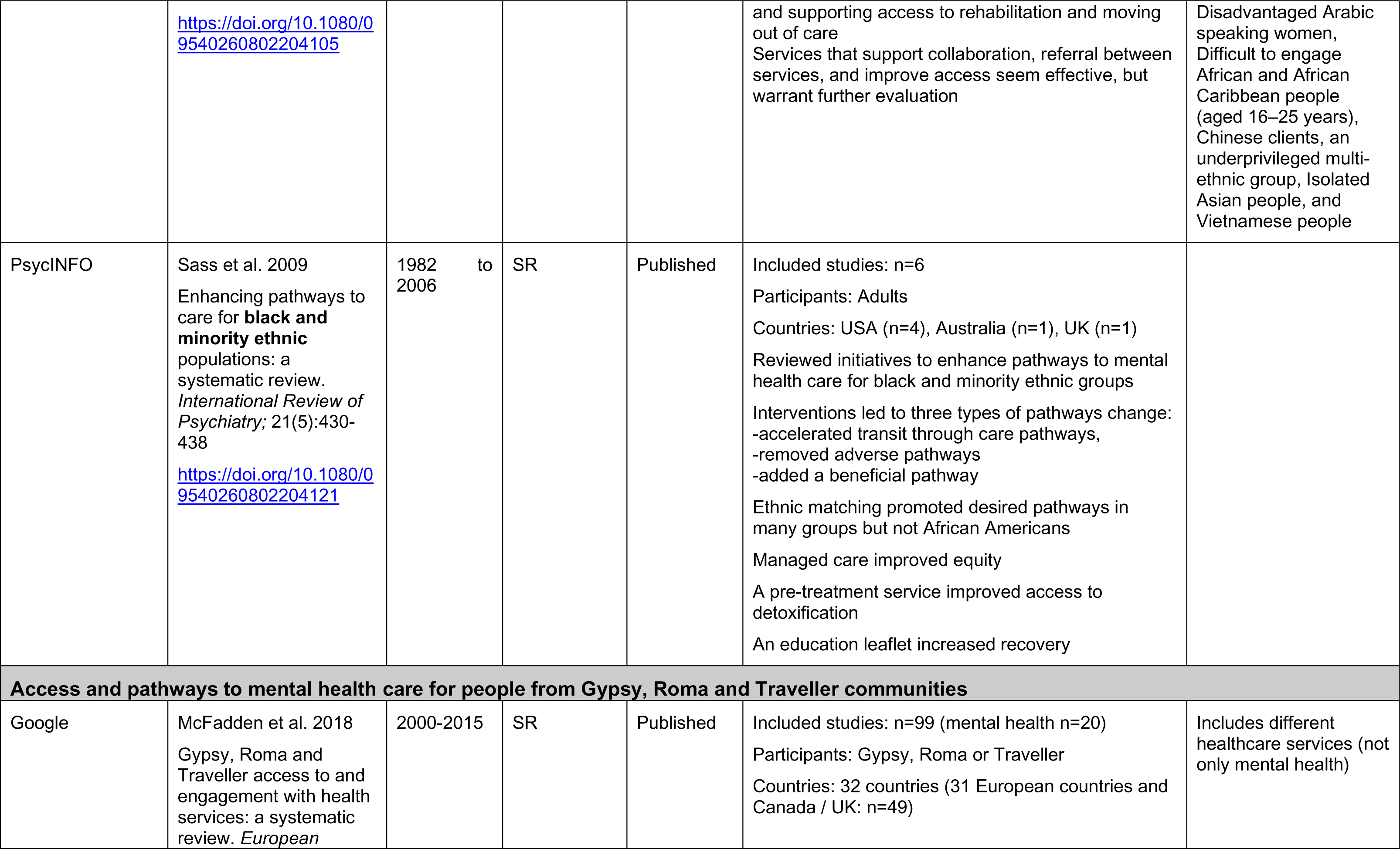
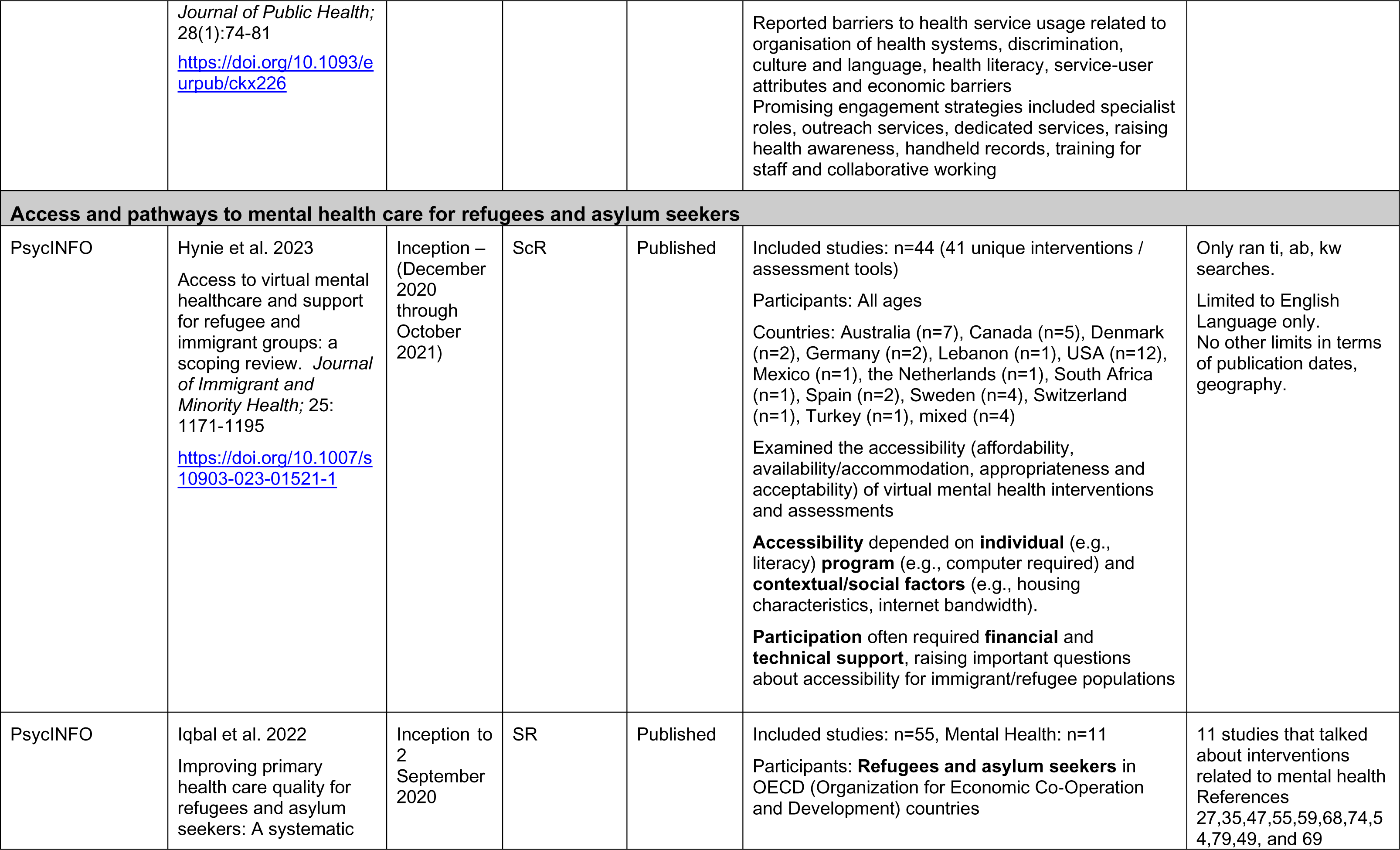
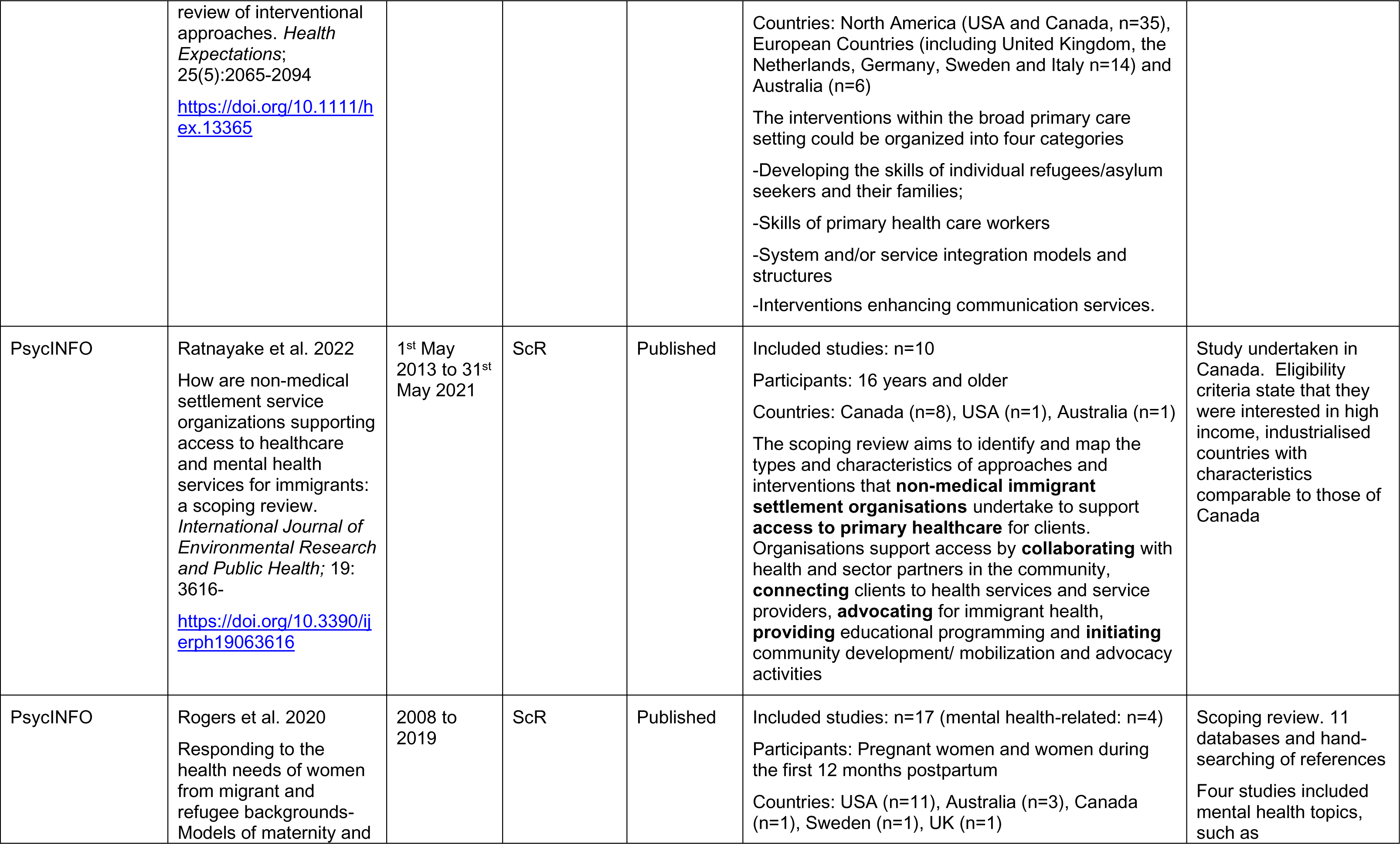
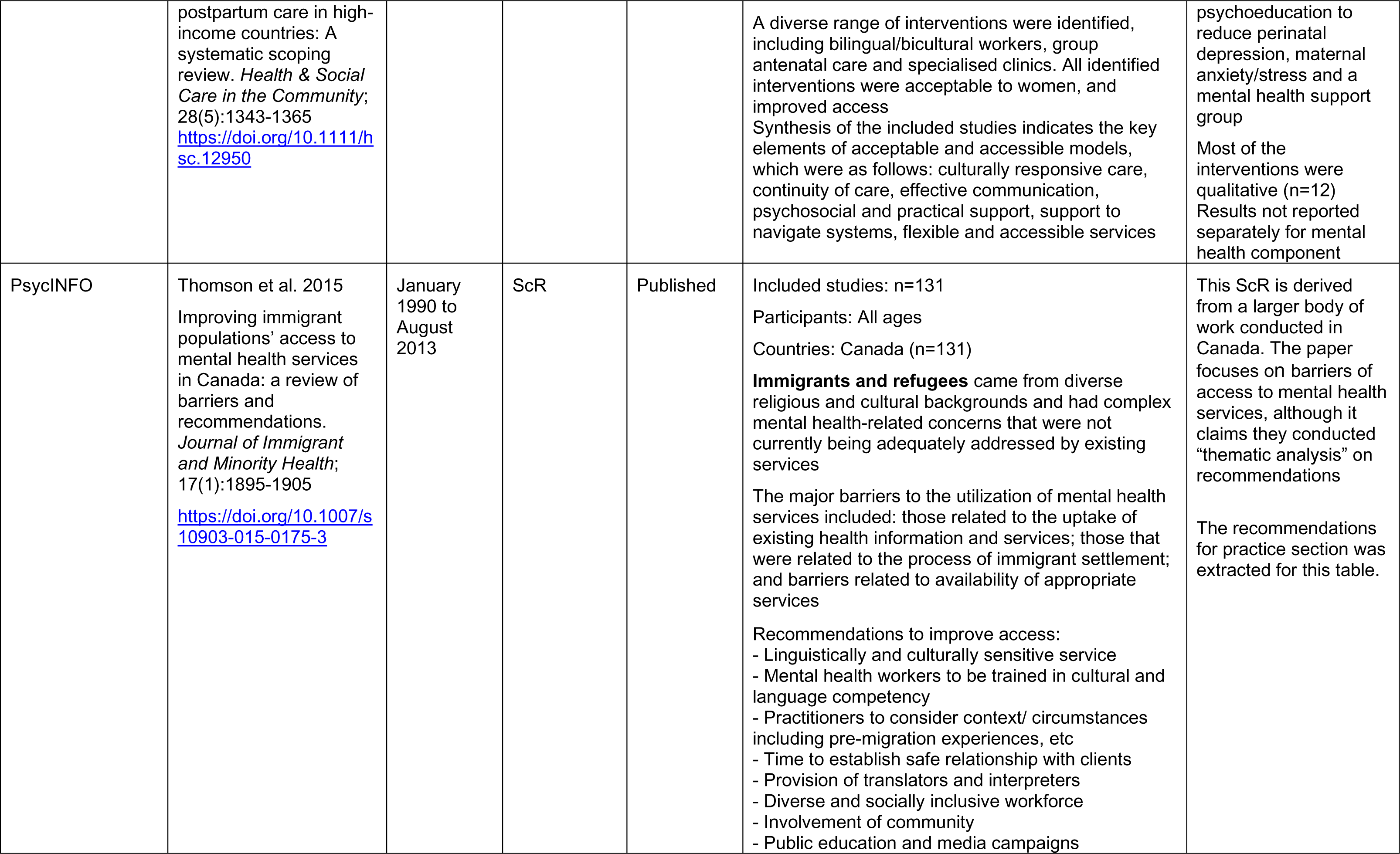
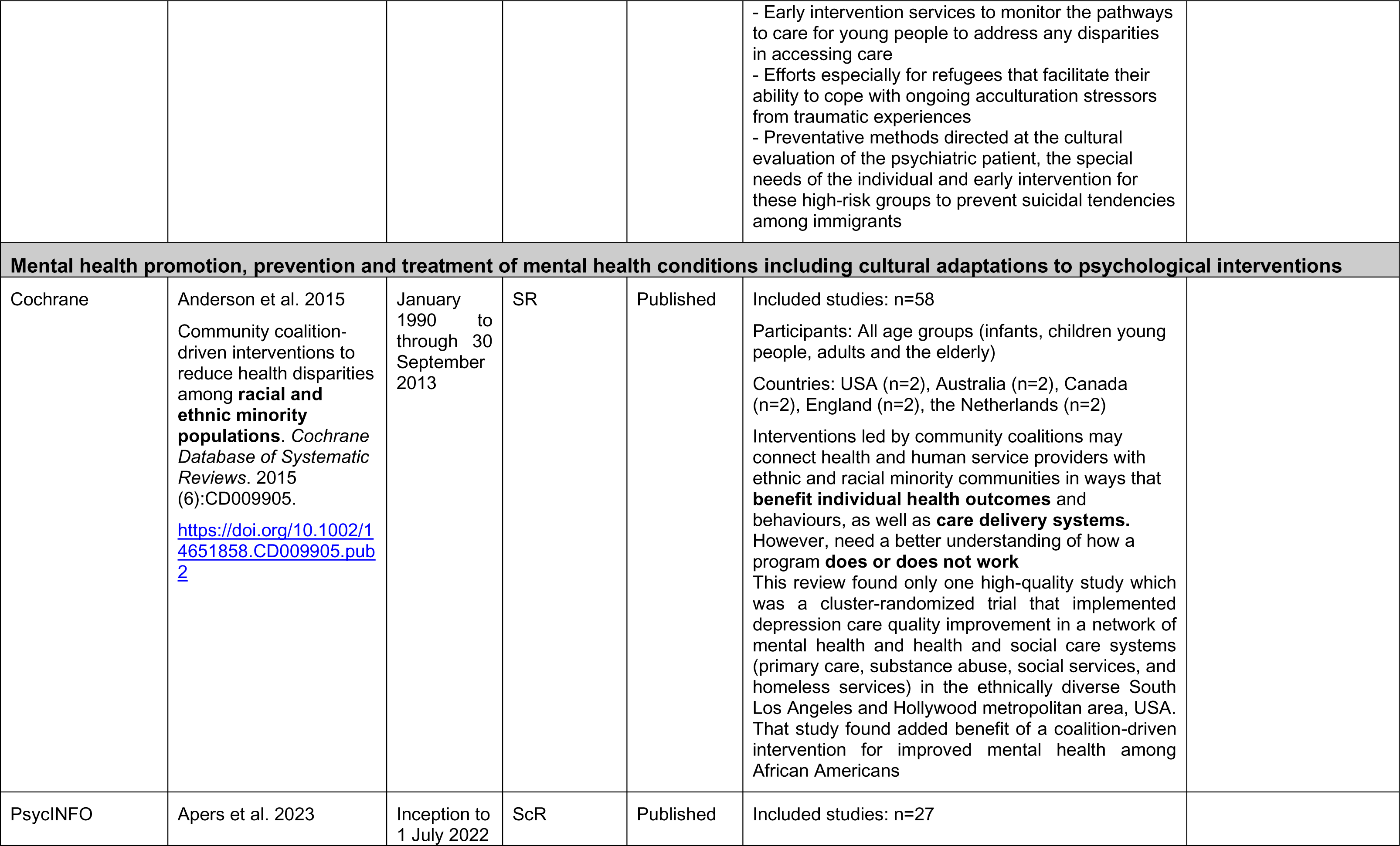
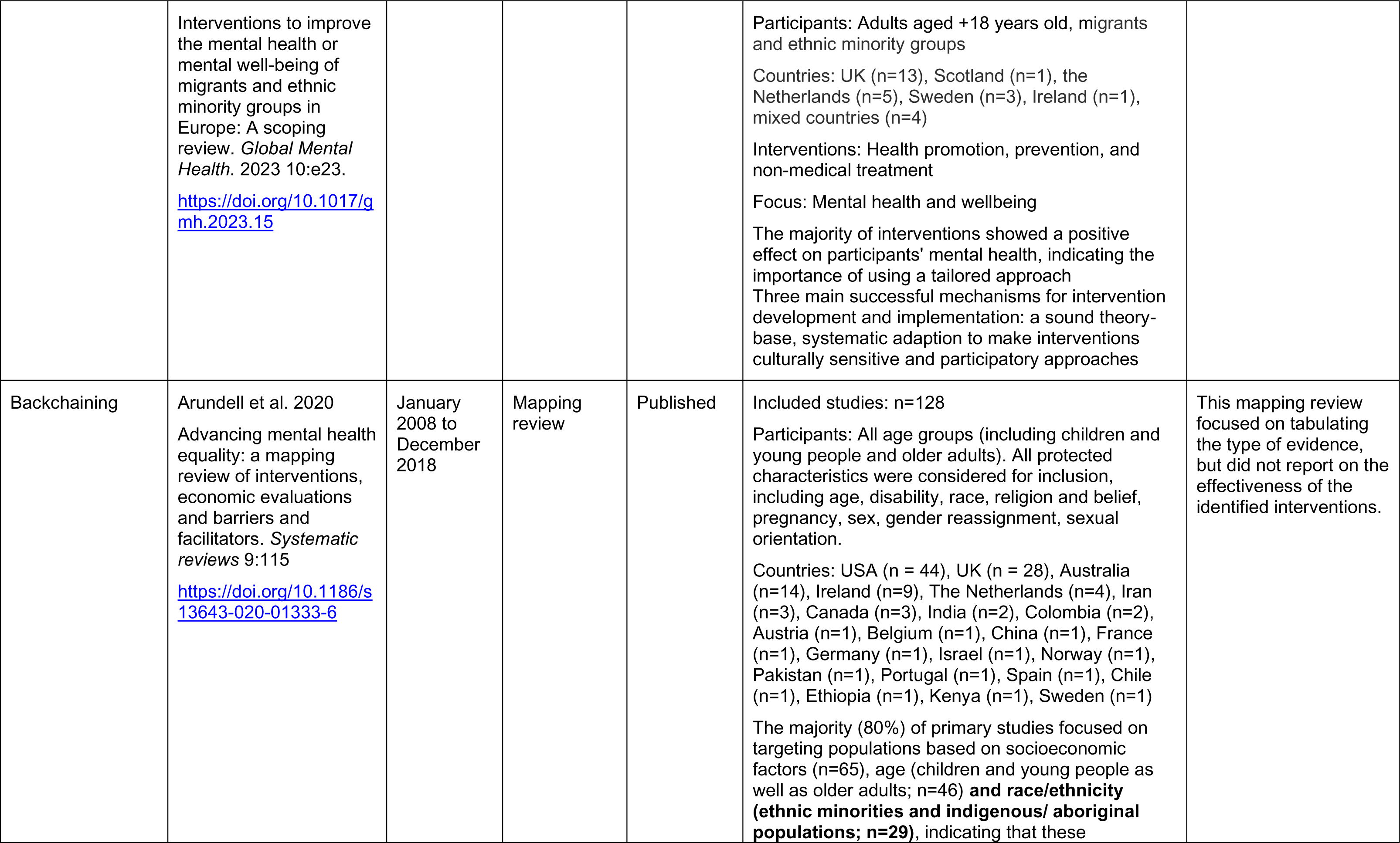
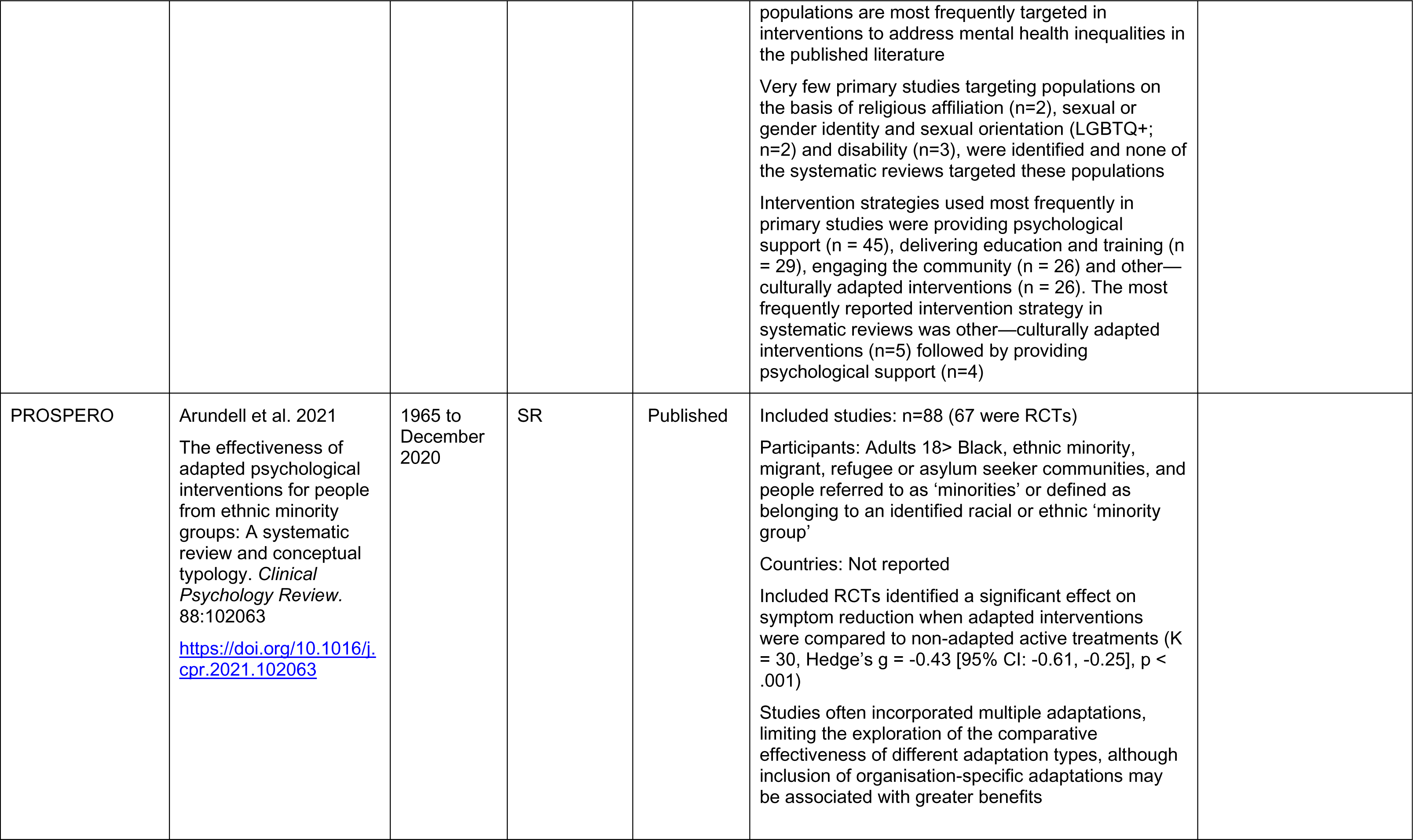
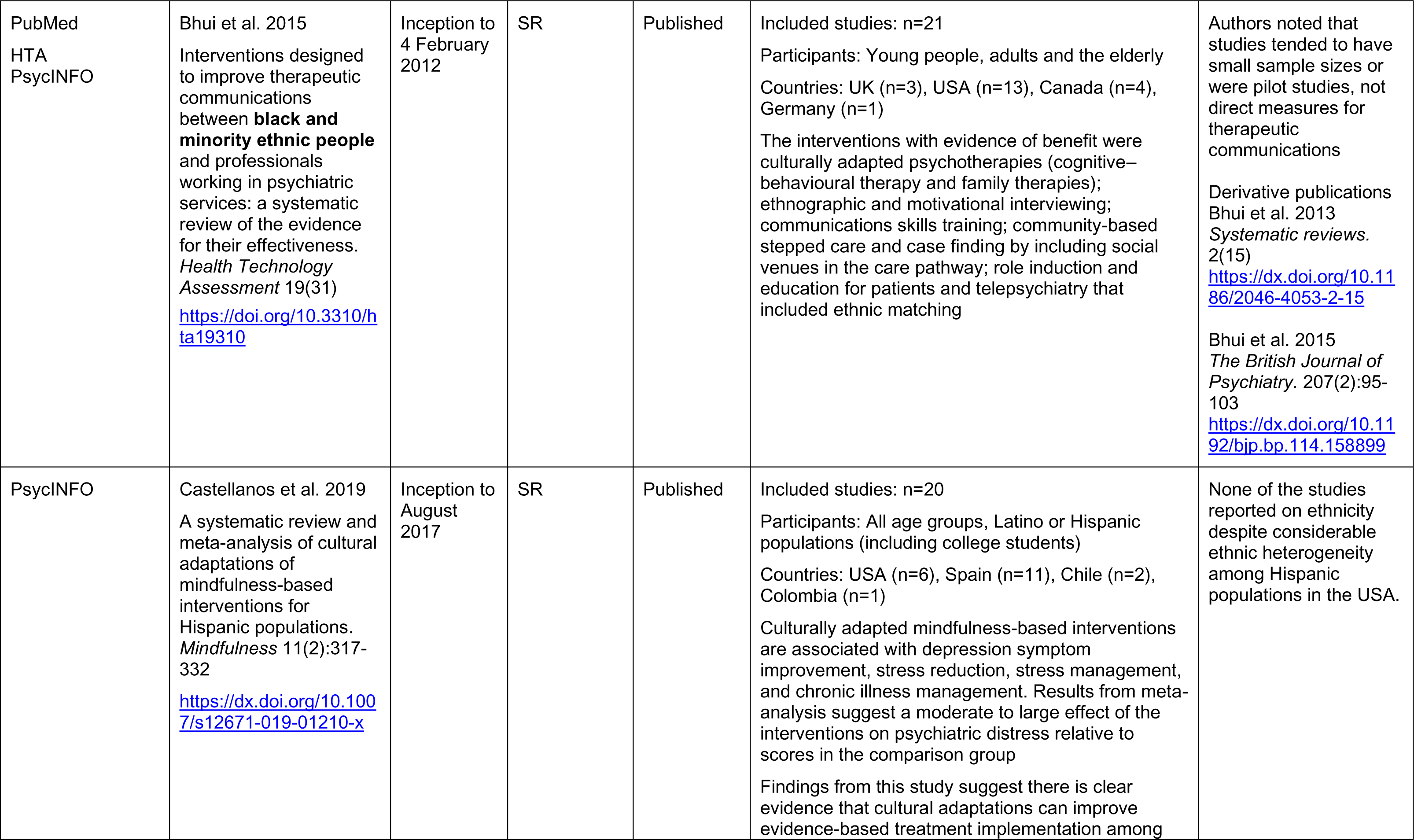
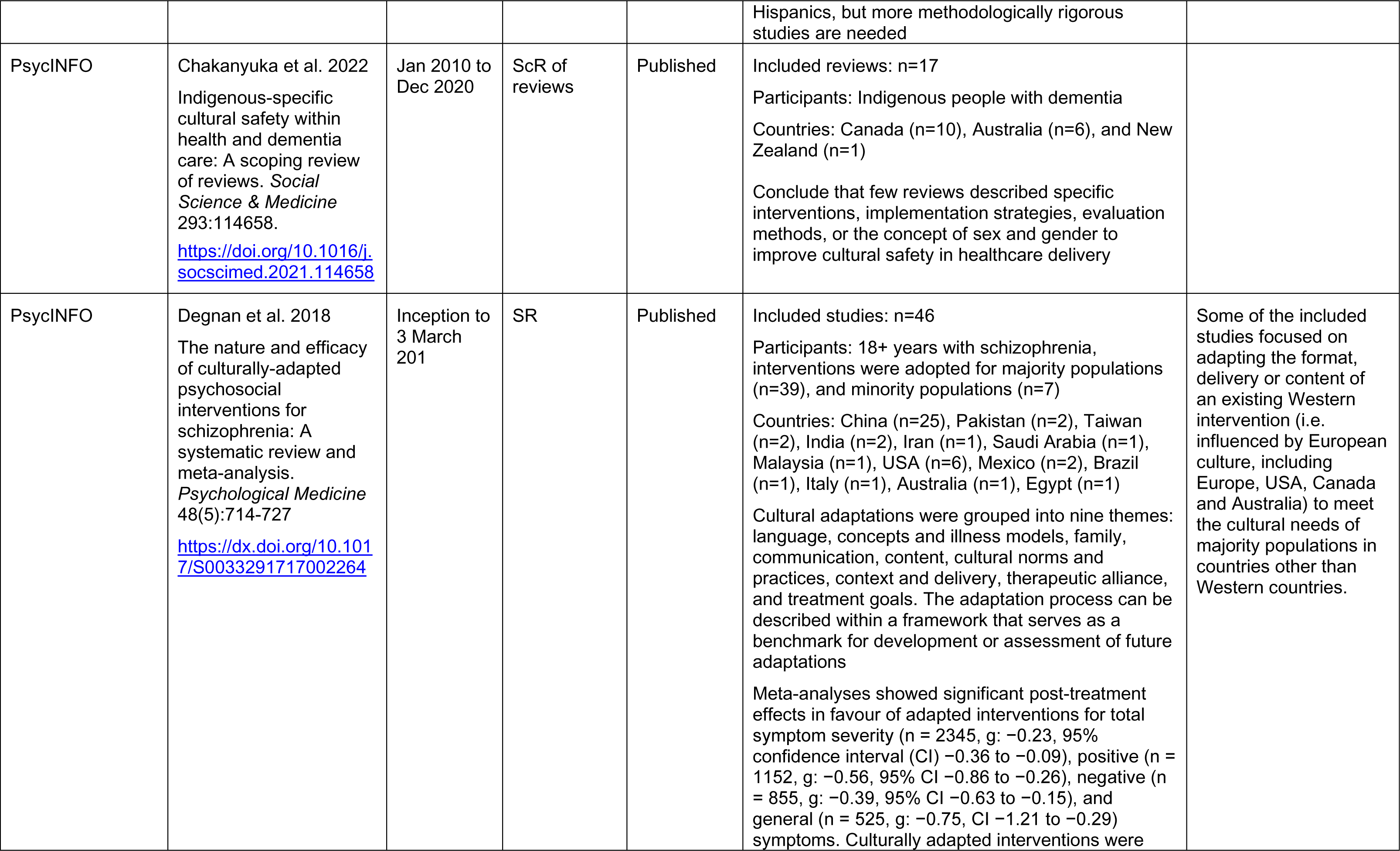
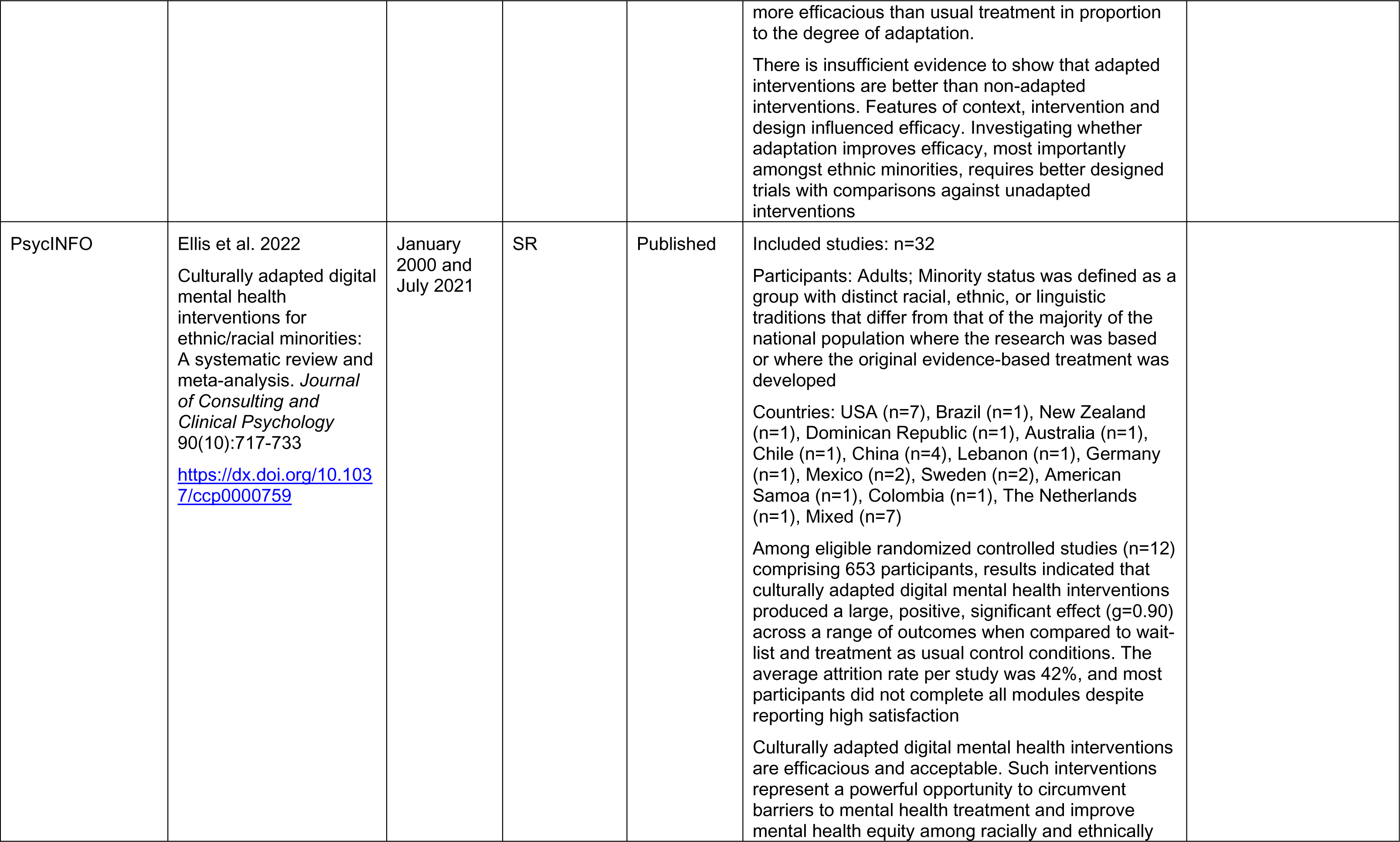
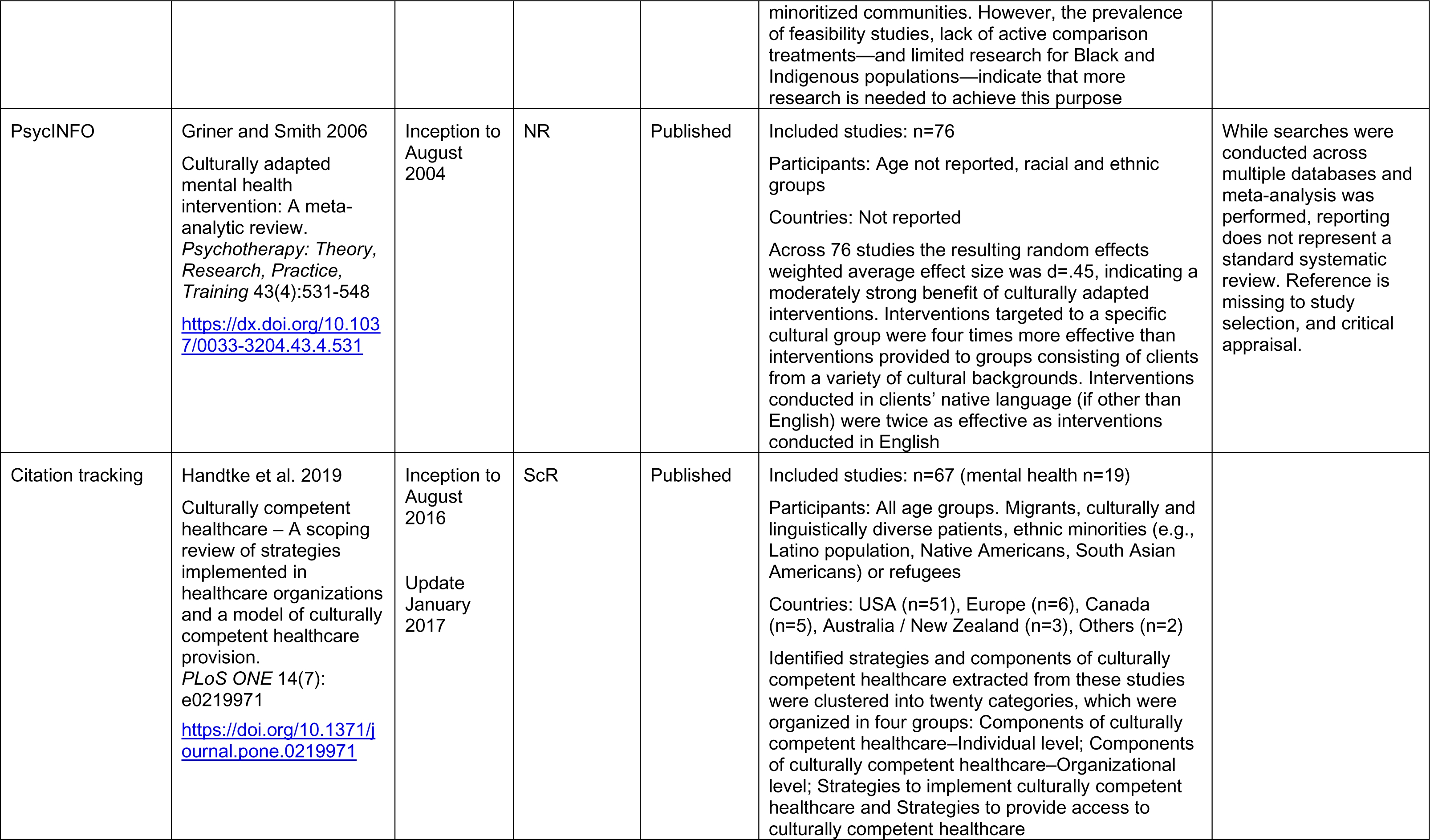
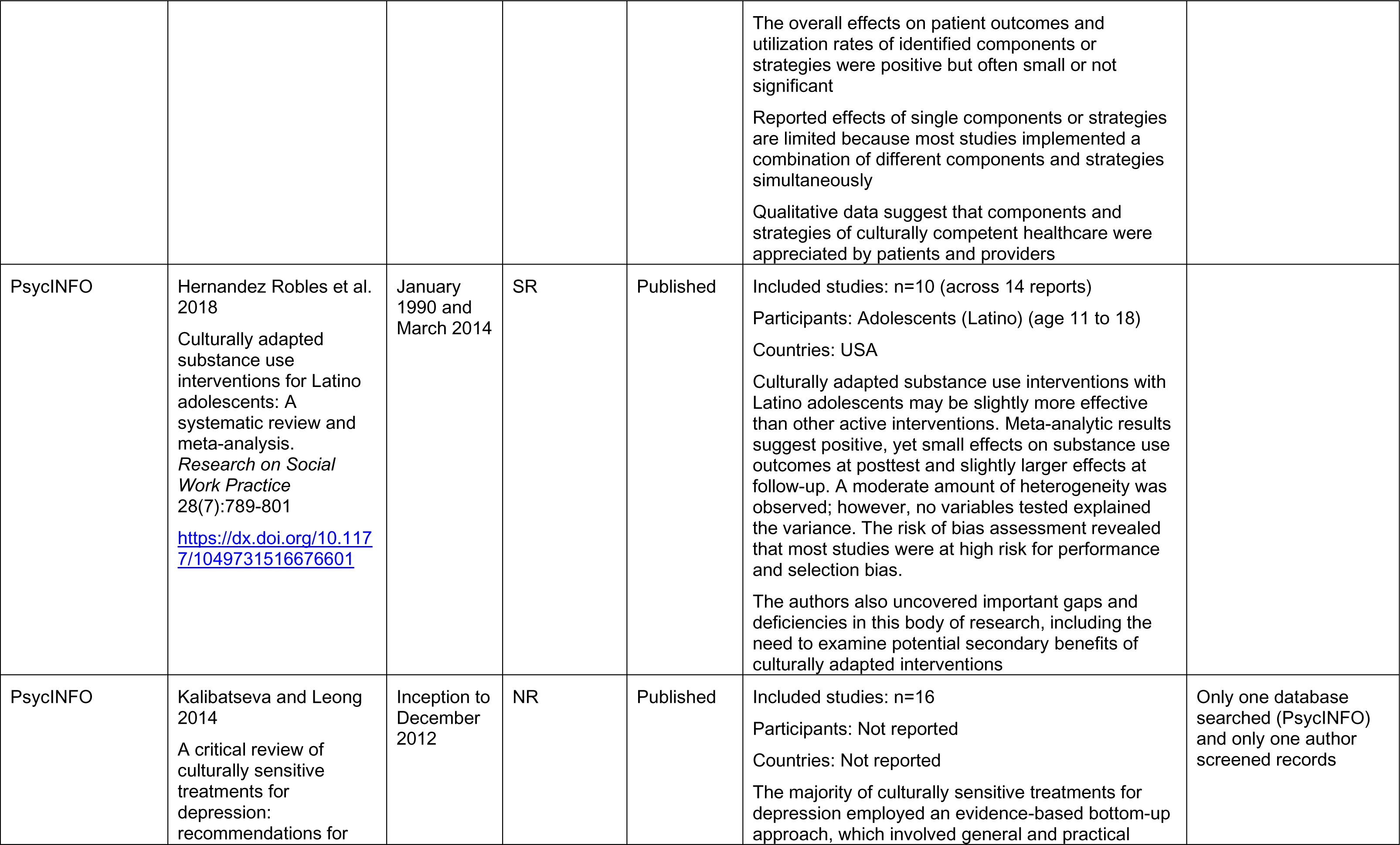
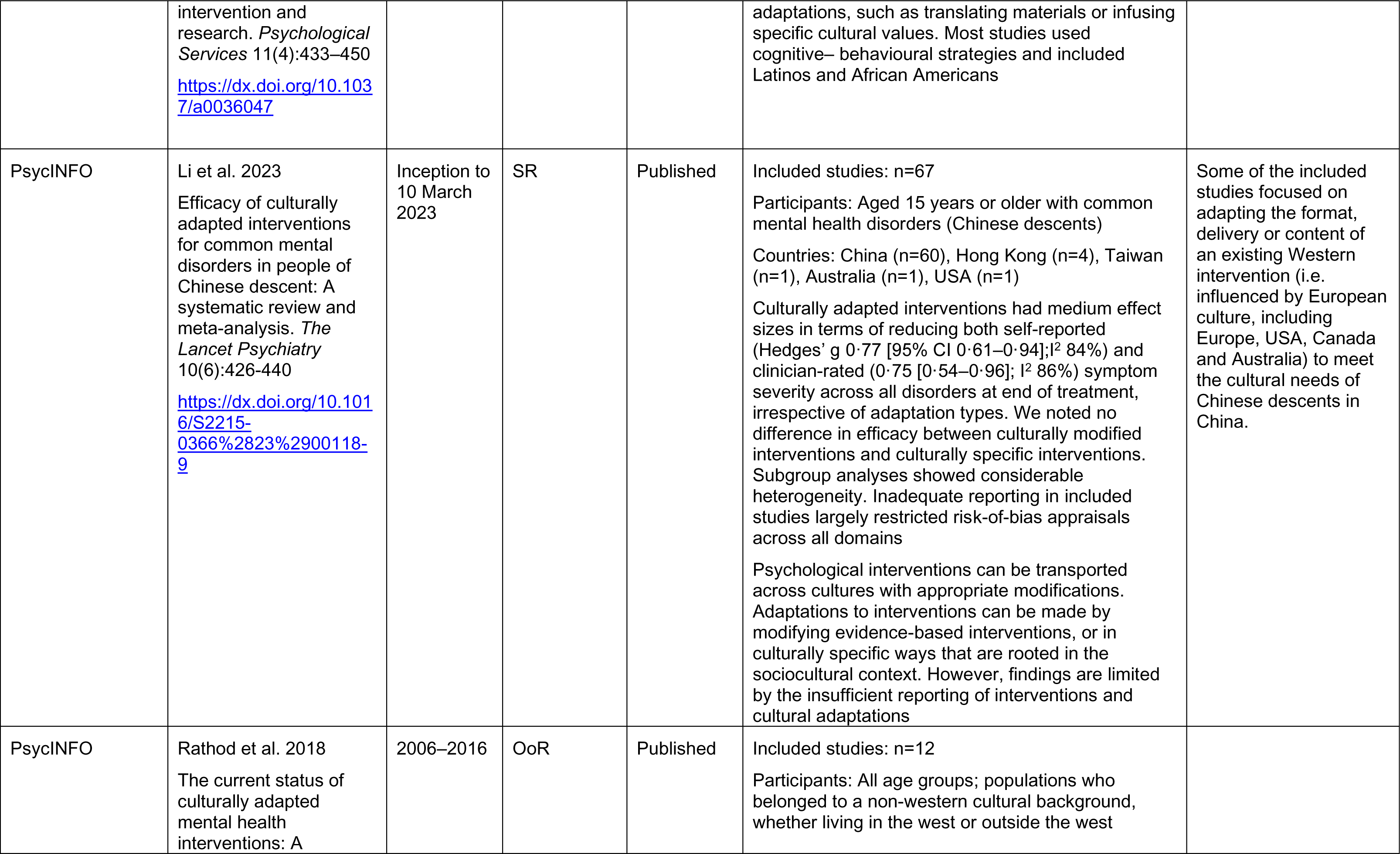
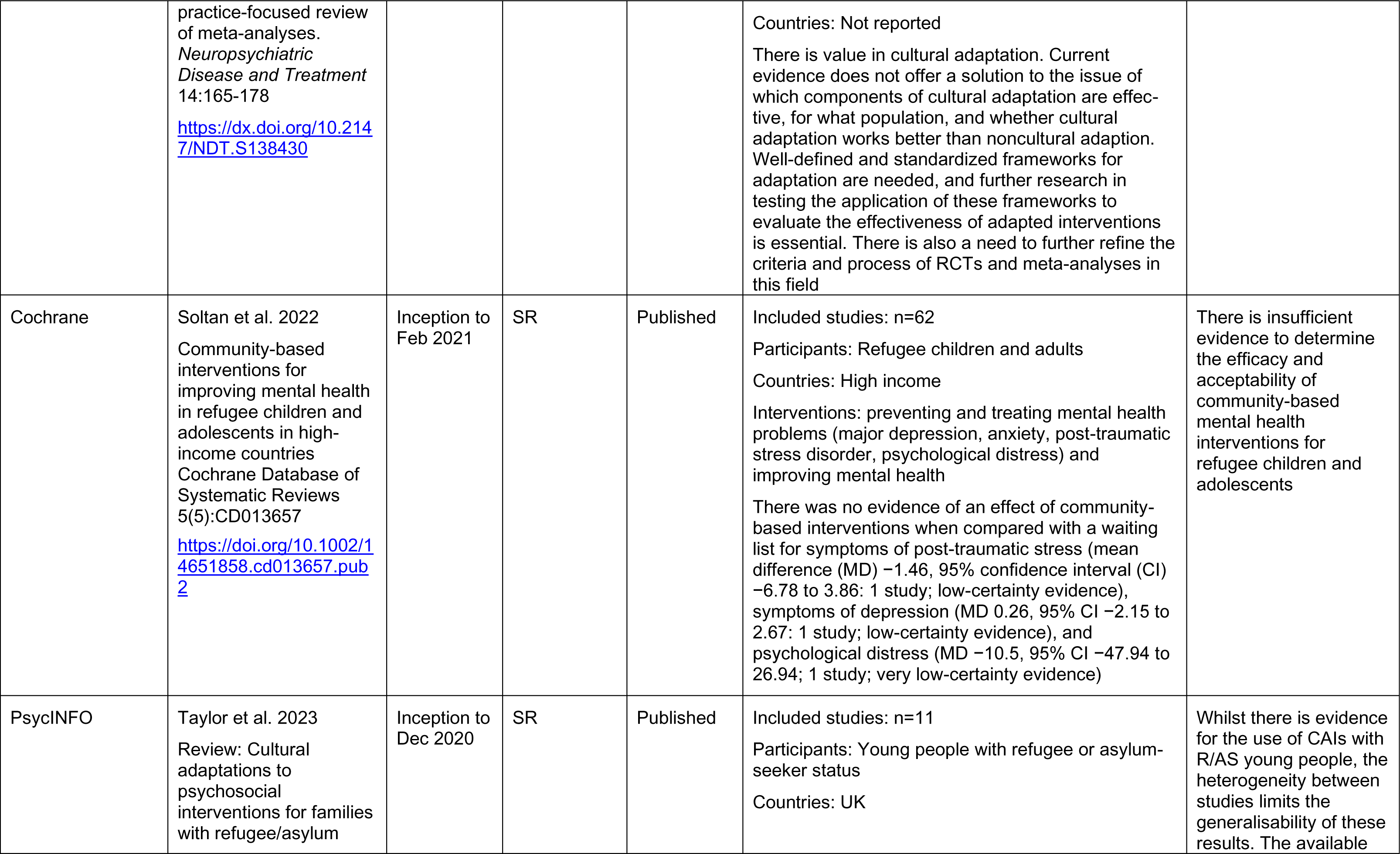
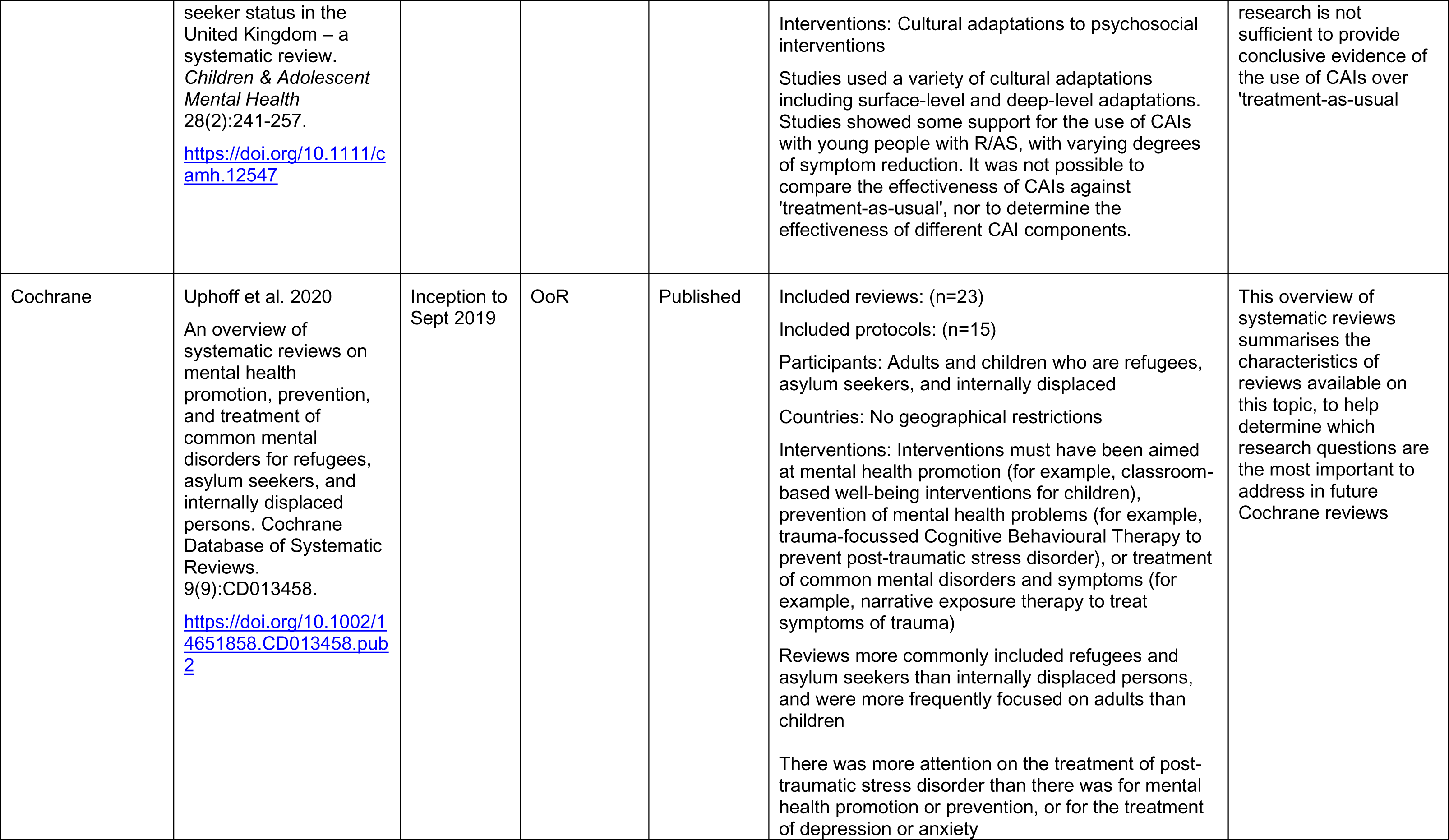
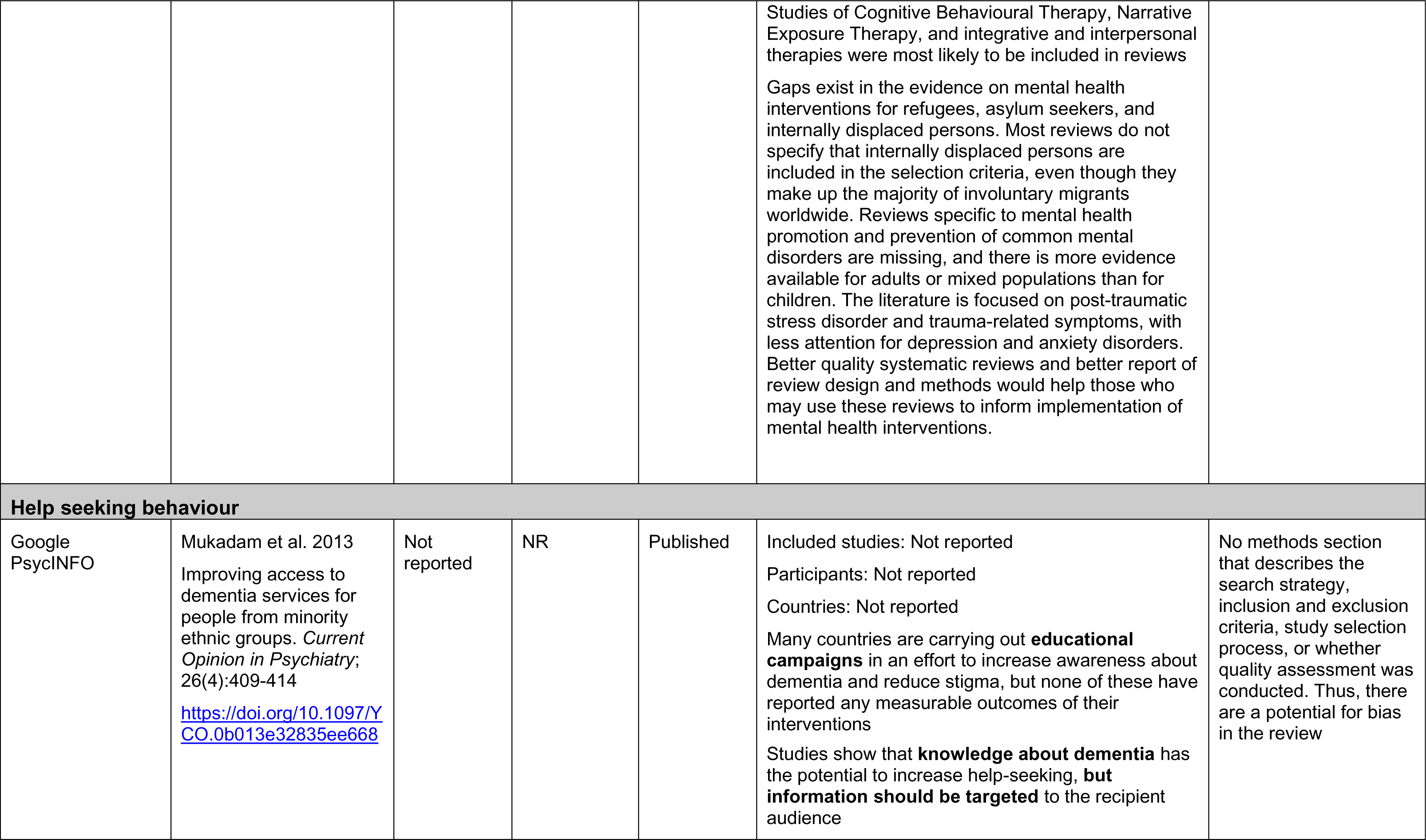
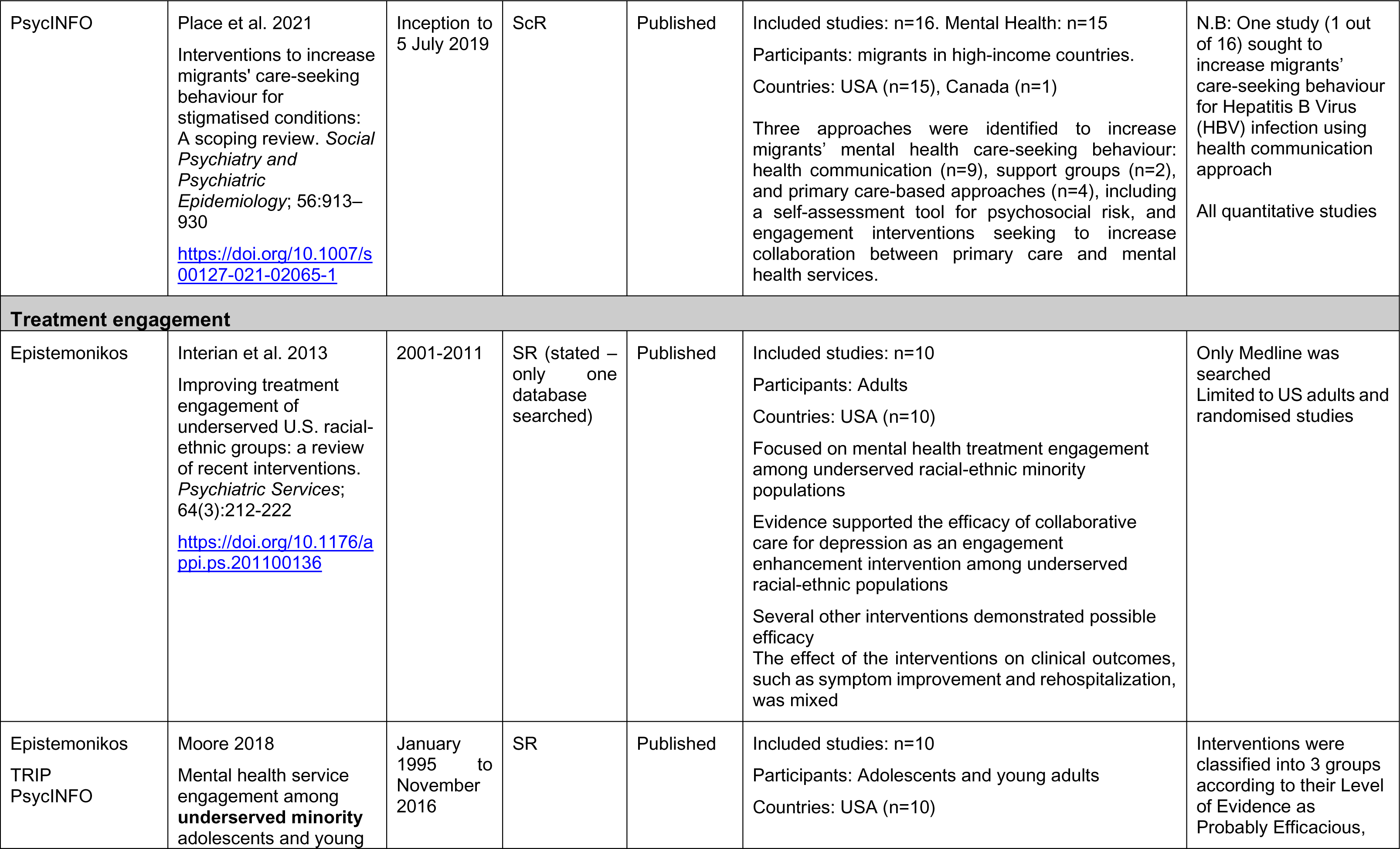
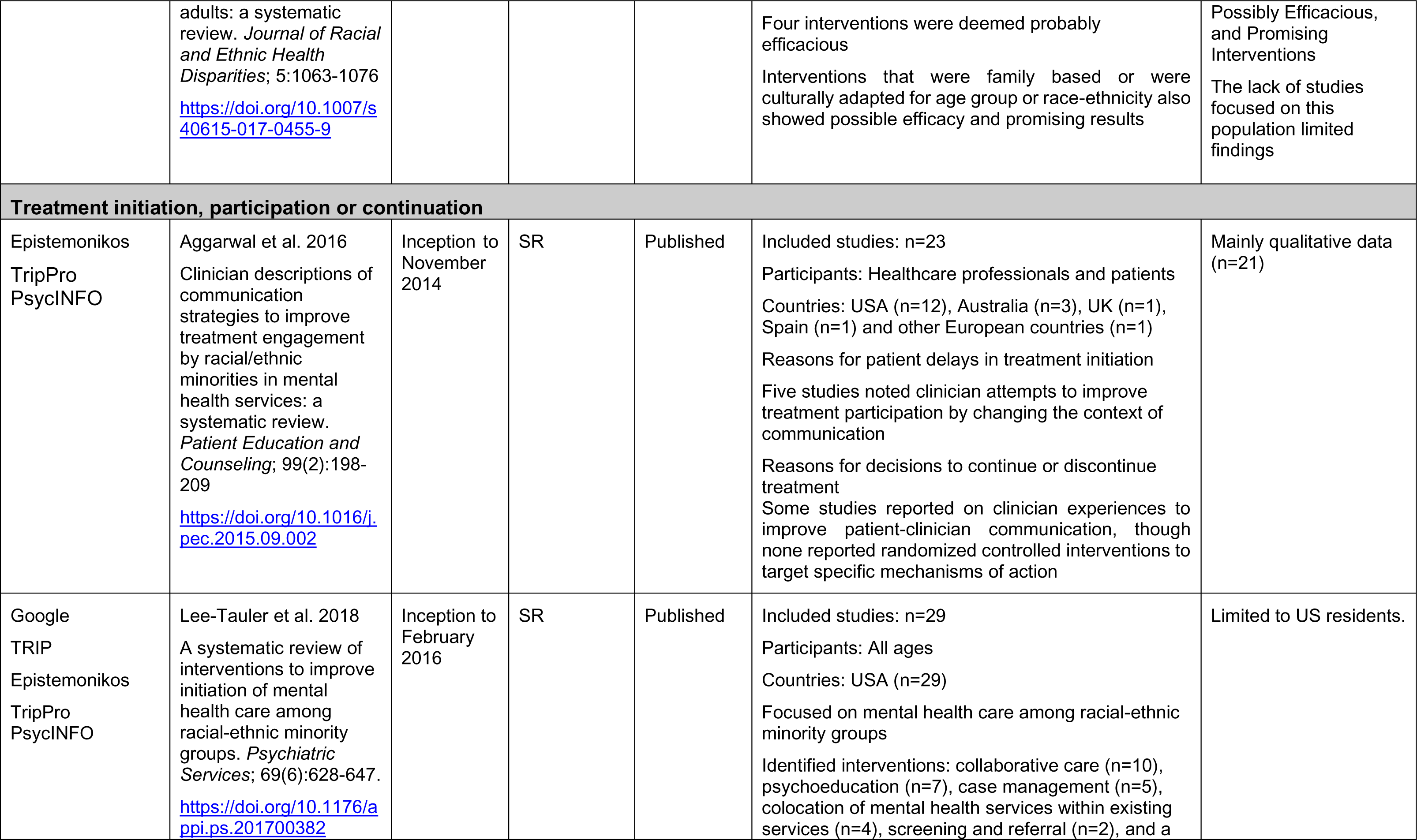
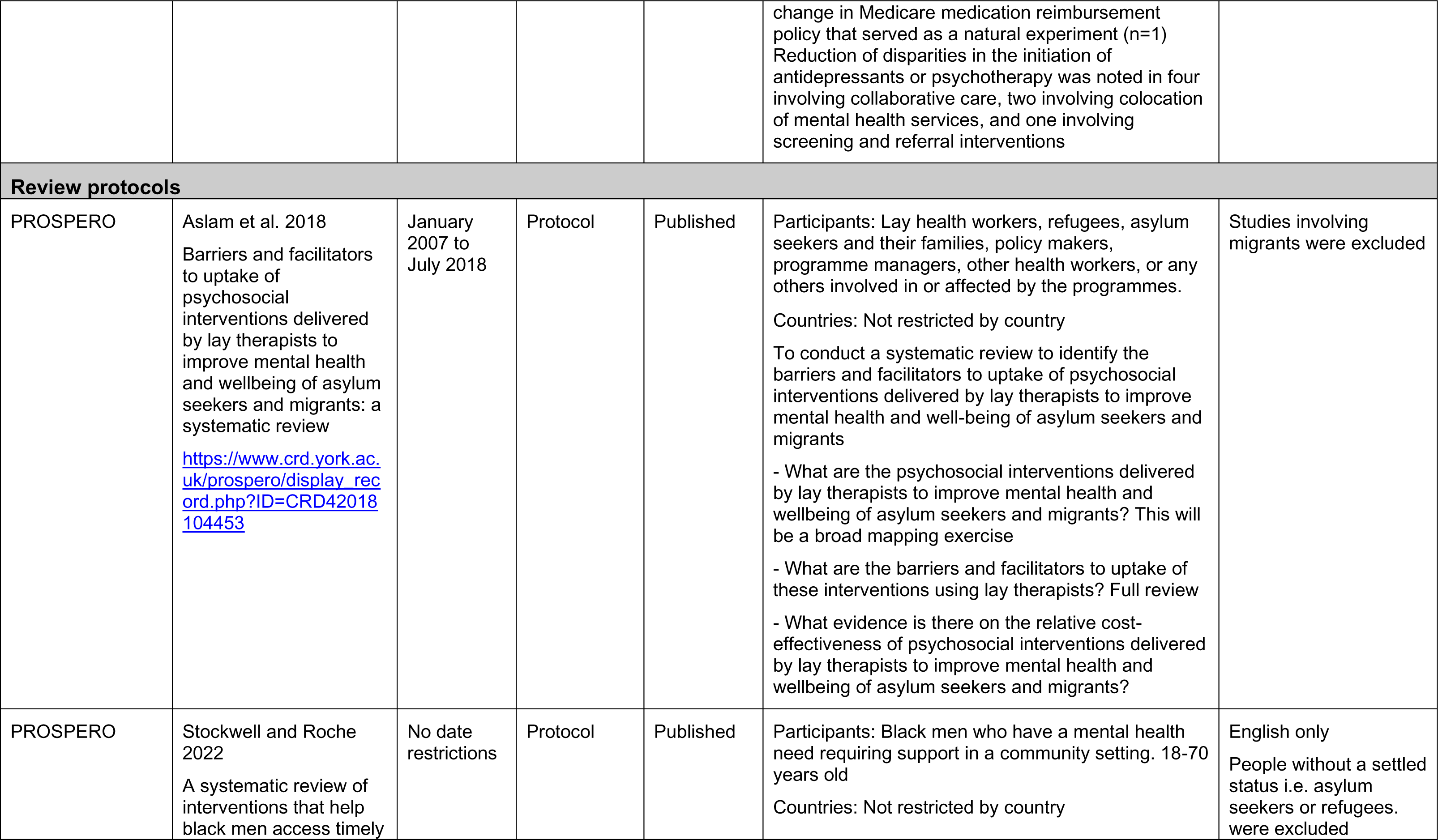
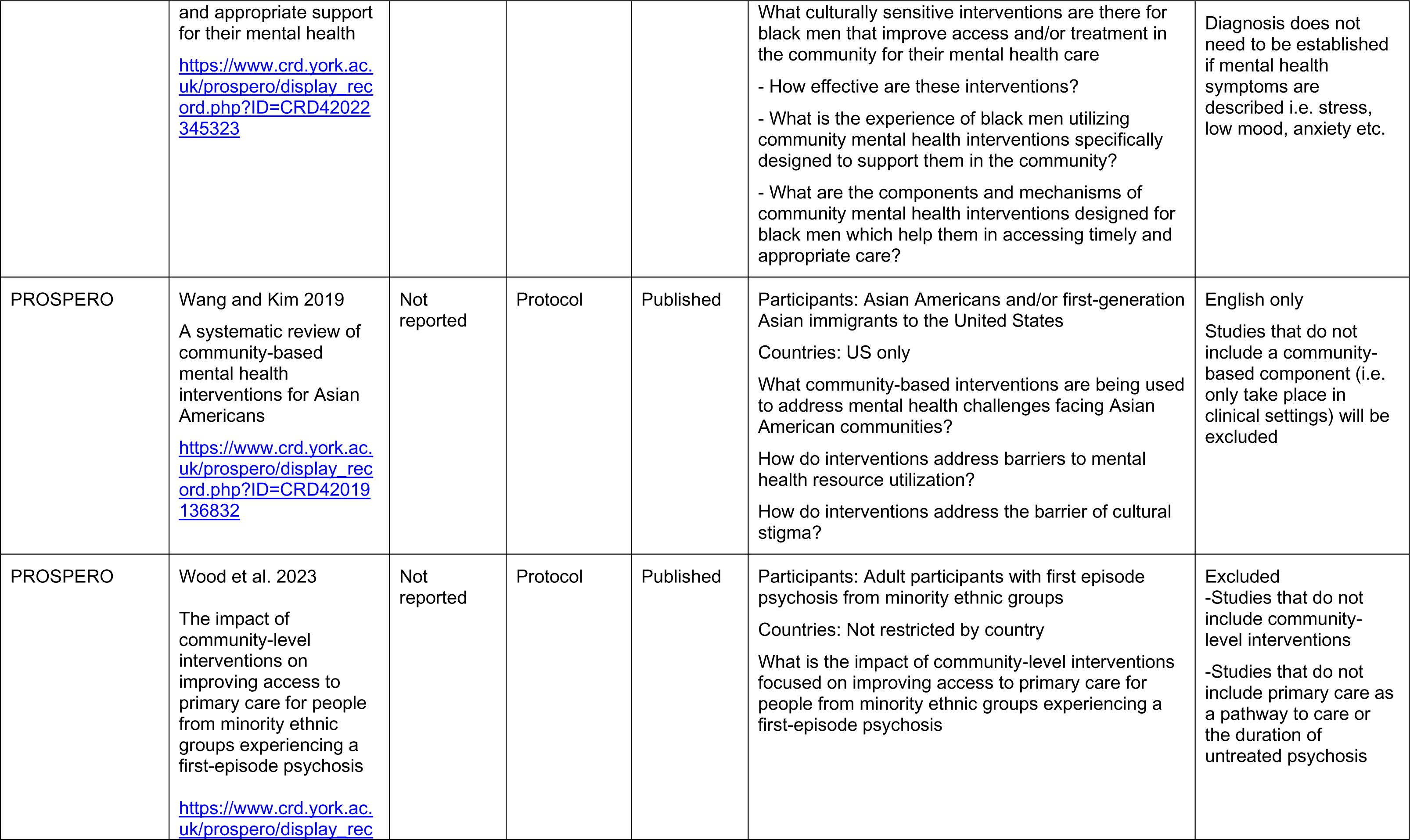
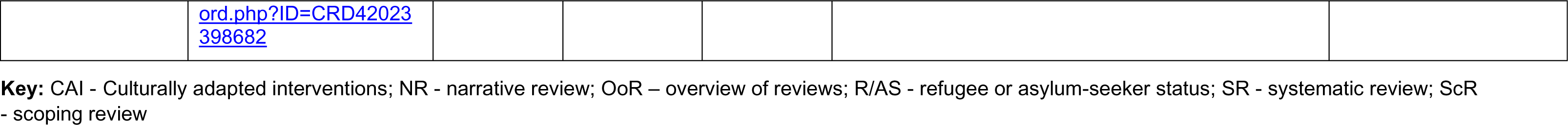
Summary of included secondary evidence.

**Table 3:**
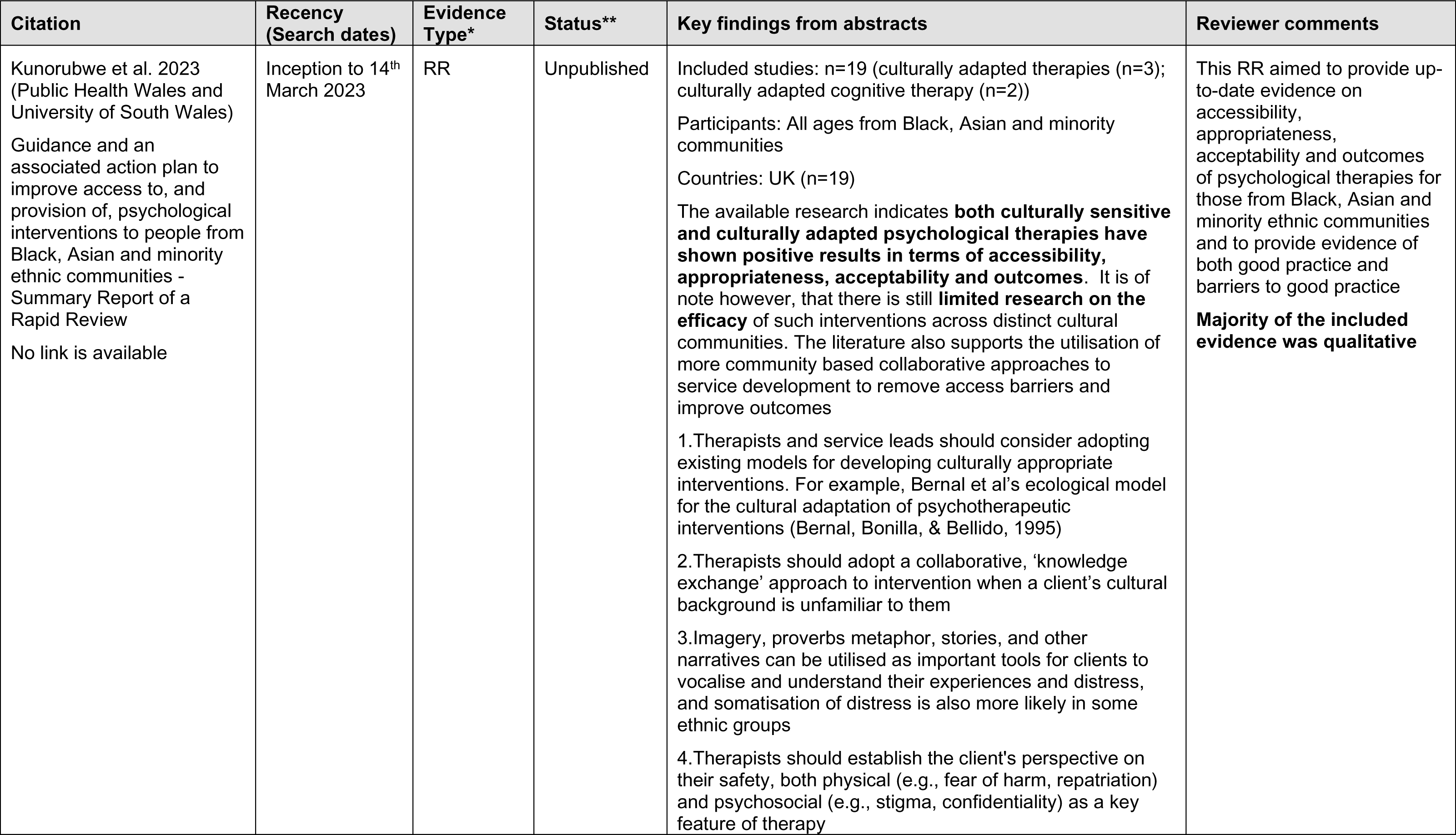
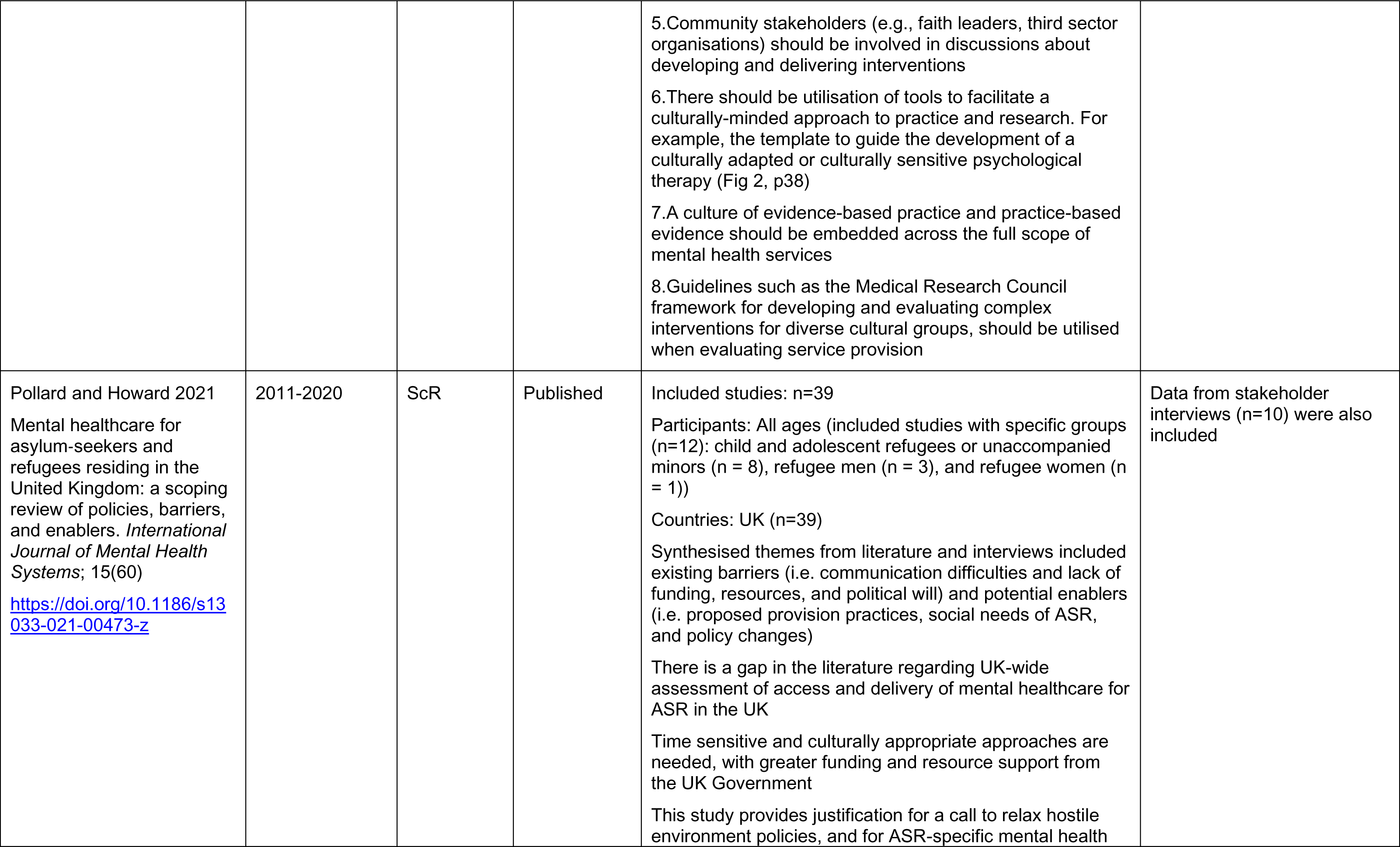
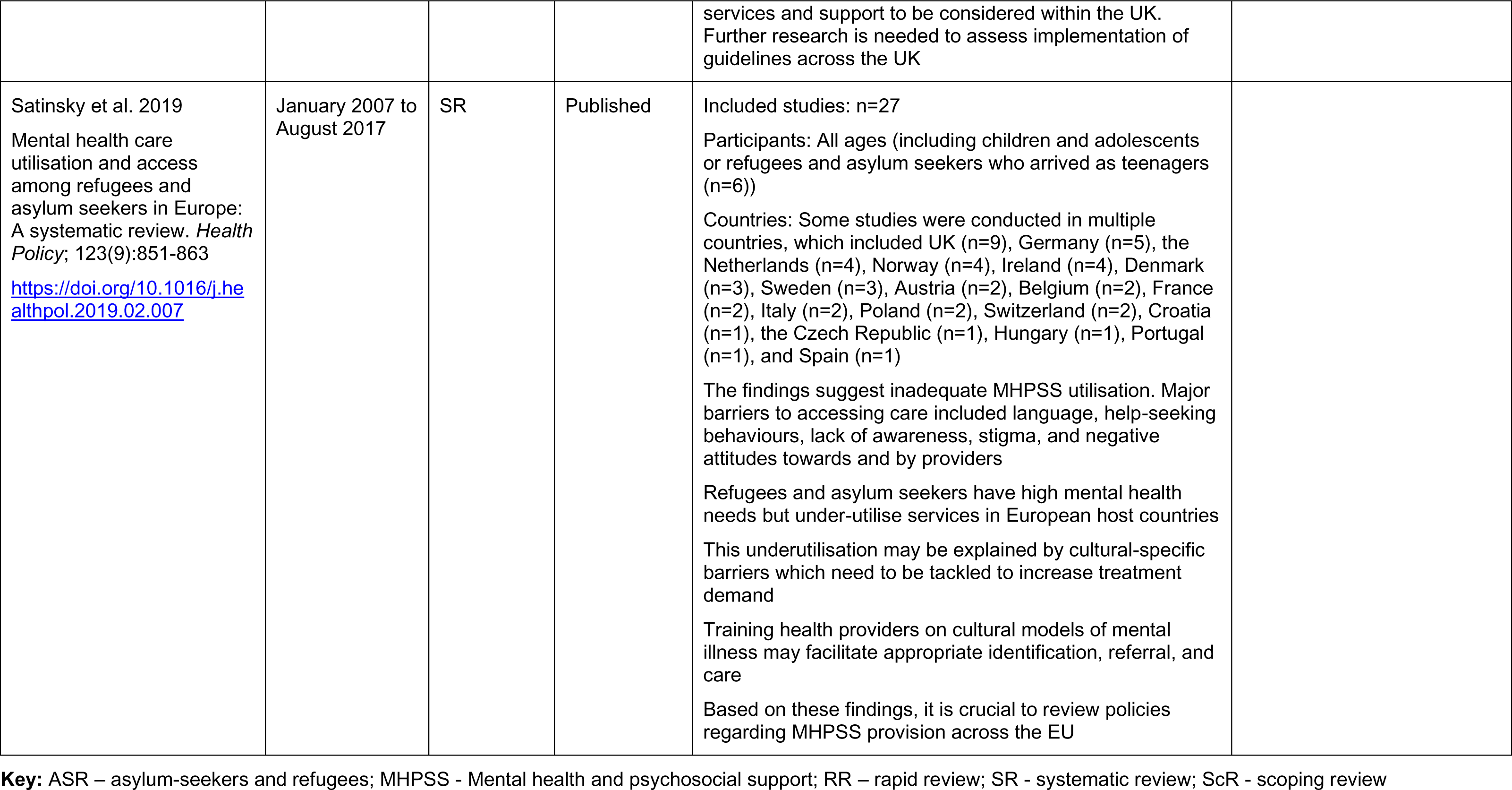
Summary of included secondary evidence identified by stakeholders.

| Citation | Recency (Search dates) | Evidence Type* | Status** | Key findings from abstracts | Reviewer comments |
| --- | --- | --- | --- | --- | --- |
| Kunorubwe et al. 2023 (Public Health Wales and University of South Wales)<br>Guidance and an associated action plan to improve access to, and provision of, psychological interventions to people from Black, Asian and minority ethnic communities - Summary Report of a Rapid Review<br>No link is available | Inception to 14<sup>th</sup> March 2023 | RR | Unpublished | Included studies: n=19 (culturally adapted therapies (n=3); culturally adapted cognitive therapy (n=2))<br>Participants: All ages from Black, Asian and minority communities<br>Countries: UK (n=19)<br>The available research indicates **both culturally sensitive and culturally adapted psychological therapies have shown positive results in terms of accessibility, appropriateness, acceptability and outcomes**. It is of note however, that there is still **limited research on the efficacy** of such interventions across distinct cultural communities. The literature also supports the utilisation of more community based collaborative approaches to service development to remove access barriers and improve outcomes<br>1. Therapists and service leads should consider adopting existing models for developing culturally appropriate interventions. For example, Bernal et al's ecological model for the cultural adaptation of psychotherapeutic interventions (Bernal, Bonilla, & Bellido, 1995)<br>2. Therapists should adopt a collaborative, 'knowledge exchange' approach to intervention when a client's cultural background is unfamiliar to them<br>3. Imagery, proverbs metaphor, stories, and other narratives can be utilised as important tools for clients to vocalise and understand their experiences and distress, and somatisation of distress is also more likely in some ethnic groups<br>4. Therapists should establish the client's perspective on their safety, both physical (e.g., fear of harm, repatriation) and psychosocial (e.g., stigma, confidentiality) as a key feature of therapy | This RR aimed to provide up-to-date evidence on accessibility, appropriateness, acceptability and outcomes of psychological therapies for those from Black, Asian and minority ethnic communities and to provide evidence of both good practice and barriers to good practice<br>**Majority of the included evidence was qualitative** |
|  |  |  |  | 5. Community stakeholders (e.g., faith leaders, third sector organisations) should be involved in discussions about developing and delivering interventions<br>6. There should be utilisation of tools to facilitate a culturally-minded approach to practice and research. For example, the template to guide the development of a culturally adapted or culturally sensitive psychological therapy (Fig 2, p38)<br>7. A culture of evidence-based practice and practice-based evidence should be embedded across the full scope of mental health services<br>8. Guidelines such as the Medical Research Council framework for developing and evaluating complex interventions for diverse cultural groups, should be utilised when evaluating service provision |  |
| Pollard and Howard 2021<br>Mental healthcare for asylum-seekers and refugees residing in the United Kingdom: a scoping review of policies, barriers, and enablers. *International Journal of Mental Health Systems*; 15(60)<br>https://doi.org/10.1186/s13033-021-00473-z | 2011-2020 | ScR | Published | Included studies: n=39<br>Participants: All ages (included studies with specific groups (n=12): child and adolescent refugees or unaccompanied minors (n = 8), refugee men (n = 3), and refugee women (n = 1))<br>Countries: UK (n=39)<br>Synthesised themes from literature and interviews included existing barriers (i.e. communication difficulties and lack of funding, resources, and political will) and potential enablers (i.e. proposed provision practices, social needs of ASR, and policy changes)<br>There is a gap in the literature regarding UK-wide assessment of access and delivery of mental healthcare for ASR in the UK<br>Time sensitive and culturally appropriate approaches are needed, with greater funding and resource support from the UK Government<br>This study provides justification for a call to relax hostile environment policies, and for ASR-specific mental health<br>services and support to be considered within the UK. Further research is needed to assess implementation of guidelines across the UK | Data from stakeholder interviews (n=10) were also included |
| Satinsky et al. 2019<br>Mental health care utilisation and access among refugees and asylum seekers in Europe: A systematic review. *Health Policy*; 123(9):851-863<br>https://doi.org/10.1016/j.healthpol.2019.02.007 | January 2007 to August 2017 | SR | Published | Included studies: n=27<br>Participants: All ages (including children and adolescents or refugees and asylum seekers who arrived as teenagers (n=6))<br>Countries: Some studies were conducted in multiple countries, which included UK (n=9), Germany (n=5), the Netherlands (n=4), Norway (n=4), Ireland (n=4), Denmark (n=3), Sweden (n=3), Austria (n=2), Belgium (n=2), France (n=2), Italy (n=2), Poland (n=2), Switzerland (n=2), Croatia (n=1), the Czech Republic (n=1), Hungary (n=1), Portugal (n=1), and Spain (n=1)<br>The findings suggest inadequate MHPSS utilisation. Major barriers to accessing care included language, help-seeking behaviours, lack of awareness, stigma, and negative attitudes towards and by providers<br>Refugees and asylum seekers have high mental health needs but under-utilise services in European host countries<br>This underutilisation may be explained by cultural-specific barriers which need to be tackled to increase treatment demand<br>Training health providers on cultural models of mental illness may facilitate appropriate identification, referral, and care<br>Based on these findings, it is crucial to review policies regarding MHPSS provision across the EU |  |

**Key:** ASR – asylum-seekers and refugees; MHPSS - Mental health and psychosocial support; RR – rapid review; SR - systematic review; ScR - scoping review

**Table 4:** Summary of included organisational reports identified by stakeholders.

| Citation<br>Citation retrieval source | Details |
| --- | --- |
| Roberts 2023<br>Psychological interventions for refugees and asylum seekers: current evidence<br>No link is available<br>Identified by stakeholder group | <u>Type of publication</u><br>Presentation (unpublished)<br>Overview of evidence that included:<br>-A network meta-analysis of psychosocial interventions for refugees and asylum seekers with PTSD https://gh.bmj.com/content/6/6/e005029.long n=23 studies<br>-Residual posttraumatic stress disorder symptoms after provision of brief behavioral intervention in low- and middle-income countries: An individual-patient data meta-analysis https://doi.org/10.1002/da.23221<br>-And 5 RCTs<br><u>Focus</u><br>To scope the evidence on the management of post-traumatic stress disorder<br>**Cognitive behavioural therapy** (SMD=-1.41; 95% CI -2.43 to -0.38) and **eye movement desensitisation and reprocessing** (SMD=-1.30; 95% CI -2.40 to -0.20) were significantly more effective than waitlist for refugees and asylum seekers.<br>There was greater symptom reduction for **Problem Management Plus** than for enhanced treatment as usual for most symptoms for refugees and asylum seekers. Hyperarousal symptoms were the most common residual symptoms after PM+, with more than 30% of participants reporting persistent sleep disturbance, concentration difficulties, and anger. |
| Traumatic Stress Wales 2023<br>Draft Asylum Seekers, Refugees and Migrants Mental Health Services Working Group Proposal<br>No link is available<br>Identified by stakeholder group | <u>Type of publication</u><br>Working group proposal (unpublished)<br><u>Focus</u><br>The proposal aims to result in a whole system approach that is integrated into current structures and initiatives, facilitates access to existing services and helps services to operate in a fully trauma-informed manner.<br>A scoping exercise identified good practice in Wales and across the UK:<br>Gwent Health Inclusion Service<br>-To address health inequalities within the Aneurin University Health Board catchment area. The service is a nurse-led specialist NHS community team which provides primary health care level services for vulnerable groups of all ages, with a specific focus on people who have difficulty in accessing standard primary care services<br>-A Social Determinants of Mental Health & Community Approach |
|  | Based in Aneurin Bevan University Health Board Child & Family Community Psychology team, places an emphasis not on “what is wrong” with someone but “what has happened” to them<br>-Cardiff and Vale Health Inclusion Service (CAVHIS)<br>CAVHIS provides health screening and urgent care for newly arrived **asylum seekers** and trafficked individuals moved to Cardiff by the Home Office<br>-Guy's & St Thomas' Health Inclusion Team, London<br>This team provides a primary health care level service for vulnerable groups (refugees, asylum seekers, homeless populations), and the refugee and asylum seeker services are across Southwark and Lambeth<br>-Solace (Yorkshire & Humber)<br>Solace is a third sector charitable organisation delivering psychotherapy and mental health support to refugees and asylum seekers across the Yorkshire and Humber region |
| VCSE Health and Wellbeing Alliance 2022<br>Barriers and Opportunities: Improving access to mental health support for refugees and people seeking asylum<br>https://www.redcross.org.uk/about-us/what-we-do/we-speak-up-for-change/vcse-health-and-wellbeing-alliance<br>Identified by stakeholder group | <u>Type of publication</u><br>Qualitative primary research report<br><u>Focus</u><br>To collaborate with people with lived experience of seeking asylum in England to design research which explores the barriers faced by refugees and people seeking asylum when trying to access support for their mental health<br>To explore what professionals working within organisations providing mental health and related support to refugees and people seeking asylum, including within the NHS, identify as barriers to accessing mental health support among this group<br>To enable people with lived experience of seeking asylum to identify opportunities for improving the access to, and provision of, mental health support for refugees and people seeking asylum in England |
| Welsh Government 2023<br>Research into “Good Access” in Community Pharmacy, NHS Dentistry and Allied Health Professional Services<br>https://www.gov.wales/research-good-access-community-pharmacy-nhs-dentistry-and-allied-health-professional-services<br>Identified by stakeholder group | <u>Type of publication</u><br>Qualitative primary research report<br><u>Focus</u><br>To identify key barriers and facilitators to accessing primary care across Community Pharmacy, NHS Dentistry and Allied Health Professionals<br>**To explore what good access means to people (general public) and the expectations of what good access looks like in Community Pharmacy, NHS Dentistry and Allied Health Professionals**<br>To explore perceptions of phone first and telephone consultations / triage |
|  | To explore perceptions of face-to-face appointments vs use of technology<br>To identify awareness of available options such as; NHS 111, Common Ailment Scheme; My health online and the NHS Wales App |

**Key:** SMD – Standardised Mean Difference; VCSE – Voluntary Community and Social Enterprise

## 8.2. Acknowledgments

The authors would like to thank Beverly Morgan, Kim Swain, Hannah Bayfield, Thomas Hoare, Olivia Gallen and Mel (Melanie) McAulay for their contributions during stakeholder meetings in guiding the focus of the review and interpretation of findings.

## 9. APPENDIX

### APPENDIX 1 Resources searched during Rapid Evidence Summary

A single list of resources has been developed for guiding and documenting the sources searched as part of a Rapid Evidence Summary. Not all resources will be searched, depending on relevancy. Some sources will be searched as part of the subsequent Rapid Review (or Rapid Evidence Map).

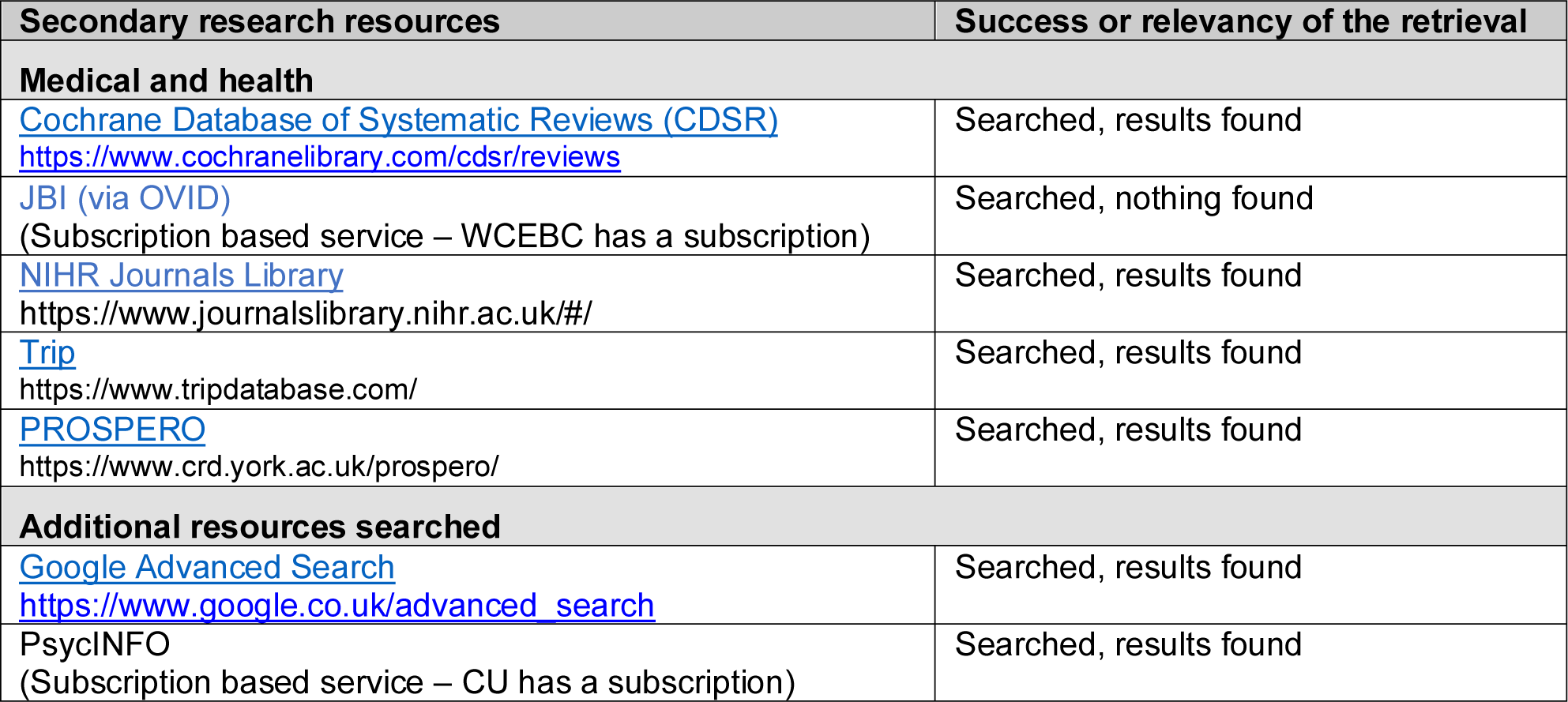

#### APA PsycInfo <1806 to November Week 1 2023> Mental health conditions

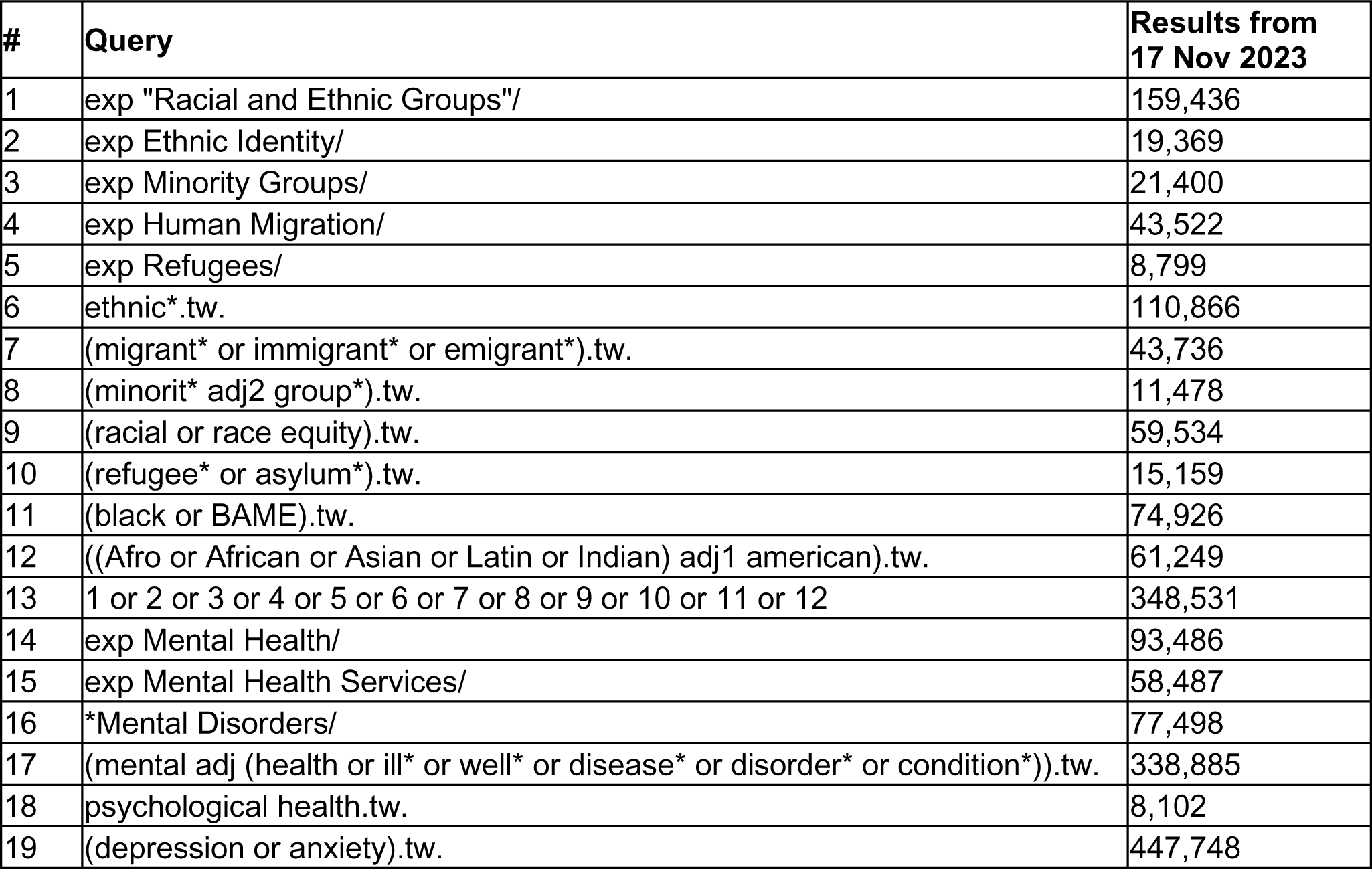
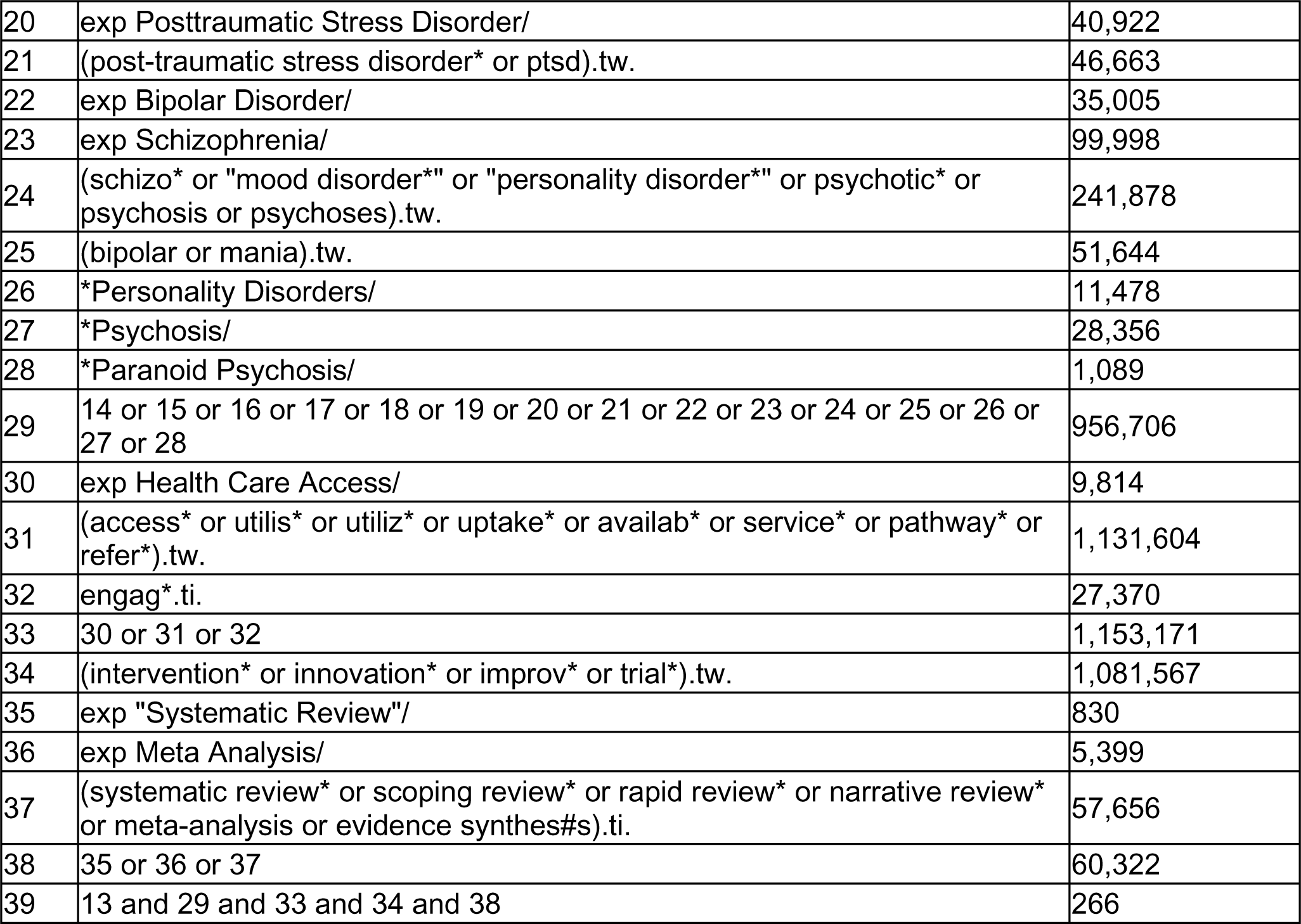

| # | Query | Results from 17 Nov 2023 |
| --- | --- | --- |
| 1 | exp "Racial and Ethnic Groups"/ | 159,436 |
| 2 | exp Ethnic Identity/ | 19,369 |
| 3 | exp Minority Groups/ | 21,400 |
| 4 | exp Human Migration/ | 43,522 |
| 5 | exp Refugees/ | 8,799 |
| 6 | ethnic*.tw. | 110,866 |
| 7 | (migrant* or immigrant* or emigrant*).tw. | 43,736 |
| 8 | (minorit* adj2 group*).tw. | 11,478 |
| 9 | (racial or race equity).tw. | 59,534 |
| 10 | (refugee* or asylum*).tw. | 15,159 |
| 11 | (black or BAME).tw. | 74,926 |
| 12 | ((Afro or African or Asian or Latin or Indian) adj1 american).tw. | 61,249 |
| 13 | 1 or 2 or 3 or 4 or 5 or 6 or 7 or 8 or 9 or 10 or 11 or 12 | 348,531 |
| 14 | exp Mental Health/ | 93,486 |
| 15 | exp Mental Health Services/ | 58,487 |
| 16 | *Mental Disorders/ | 77,498 |
| 17 | (mental adj (health or ill* or well* or disease* or disorder* or condition*)).tw. | 338,885 |
| 18 | psychological health.tw. | 8,102 |
| 19 | (depression or anxiety).tw. | 447,748 |
| 20 | exp Posttraumatic Stress Disorder/ | 40,922 |
| 21 | (post-traumatic stress disorder* or ptsd).tw. | 46,663 |
| 22 | exp Bipolar Disorder/ | 35,005 |
| 23 | exp Schizophrenia/ | 99,998 |
| 24 | (schizo* or "mood disorder*" or "personality disorder*" or psychotic* or psychosis or psychoses).tw. | 241,878 |
| 25 | (bipolar or mania).tw. | 51,644 |
| 26 | *Personality Disorders/ | 11,478 |
| 27 | *Psychosis/ | 28,356 |
| 28 | *Paranoid Psychosis/ | 1,089 |
| 29 | 14 or 15 or 16 or 17 or 18 or 19 or 20 or 21 or 22 or 23 or 24 or 25 or 26 or 27 or 28 | 956,706 |
| 30 | exp Health Care Access/ | 9,814 |
| 31 | (access* or utilis* or utiliz* or uptake* or availab* or service* or pathway* or refer*).tw. | 1,131,604 |
| 32 | engag*.ti. | 27,370 |
| 33 | 30 or 31 or 32 | 1,153,171 |
| 34 | (intervention* or innovation* or improv* or trial*).tw. | 1,081,567 |
| 35 | exp "Systematic Review"/ | 830 |
| 36 | exp Meta Analysis/ | 5,399 |
| 37 | (systematic review* or scoping review* or rapid review* or narrative review* or meta-analysis or evidence synthes#s).ti. | 57,656 |
| 38 | 35 or 36 or 37 | 60,322 |
| 39 | 13 and 29 and 33 and 34 and 38 | 266 |

#### APA PsycInfo <1806 to 28 November 2023> Dementia

| # | Query | Results from<br>28 Nov 2023 |
| --- | --- | --- |
| 1 | exp "Racial and Ethnic Groups"/ | 159,567 |
| 2 | exp Ethnic Identity/ | 19,386 |
| 3 | exp Minority Groups/ | 21,426 |
| 4 | exp Human Migration/ | 43,555 |
| 5 | exp Refugees/ | 8,814 |
| 6 | ethnic*.tw. | 110,964 |
| 7 | (migrant* or immigrant* or emigrant*).tw. | 43,764 |
| 8 | (minorit* adj2 group*).tw. | 11,486 |
| 9 | (racial or race equity).tw. | 59,594 |
| 10 | (refugee* or asylum*).tw. | 15,178 |
| 11 | (black or BAME).tw. | 75,006 |
| 12 | ((Afro or African or Asian or Latin or Indian) adj1 american).tw. | 61,282 |
| 13 | 1 or 2 or 3 or 4 or 5 or 6 or 7 or 8 or 9 or 10 or 11 or 12 | 348,828 |
| 14 | exp dementia/ | 92,970 |
| 15 | exp Alzheimer's Disease/ | 56,227 |
| 16 | dementia.tw. | 79,129 |
| 17 | alzheimer.tw. | 16,210 |
| 18 | 14 or 15 or 16 or 17 | 113,897 |
| 19 | exp Health Care Access/ | 9,838 |
| 20 | (access* or utilis* or utiliz* or uptake* or availab* or service* or pathway* or refer*).tw. | 1,132,640 |
| 21 | engag*.ti. | 27,399 |
| 22 | 19 or 20 or 21 | 1,154,227 |
| 23 | (intervention* or innovation* or improv* or trial*).tw. | 1,082,671 |
| 24 | exp "Systematic Review"/ | 832 |
| 25 | exp Meta Analysis/ | 5,401 |
| 26 | (systematic review* or scoping review* or rapid review* or narrative review* or meta-analysis or evidence synthes#s).ti. | 57,864 |
| 27 | 23 or 24 or 25 or 26 | 60,531 |
| 28 | 13 and 18 and 22 and 23 and 27 | 19 |

#### APA PsycInfo <1806 to November Week 3 2023> Gypsy, Roma or Travellers

| # | Search | Results from<br>29 Nov 2023 |
| --- | --- | --- |
| 1 | exp Romanies/ | 419 |
| 2 | roma.mp. | 865 |
| 3 | romany.mp. | 25 |
| 4 | romani.mp. | 167 |
| 5 | gypsy.mp. | 325 |
| 6 | gypsies.mp. | 293 |
| 7 | gipsies.mp. | 6 |
| 8 | travel?er*.mp. | 2,640 |
| 9 | 1 or 2 or 3 or 4 or 5 or 6 or 7 or 8 | 3,899 |
| 10 | exp Mental Health/ | 93,835 |
| 11 | exp Mental Health Services/ | 58,584 |
| 12 | *Mental Disorders/ | 77,635 |
| 13 | (mental adj (health or ill* or well* or disease* or disorder* or condition*)).tw. | 339,778 |
| 14 | psychological health.tw. | 8,123 |
| 15 | (depression or anxiety).tw. | 448,503 |
| 16 | exp Posttraumatic Stress Disorder/ | 41,013 |
| 17 | (post-traumatic stress disorder* or ptsd).tw. | 46,752 |
| 18 | exp Bipolar Disorder/ | 35,034 |
| 19 | exp Schizophrenia/ | 100,063 |
| 20 | (schizo* or "mood disorder*" or "personality disorder*" or psychotic* or psychosis or psychoses).tw. | 242,071 |
| 21 | (bipolar or mania).tw. | 51,694 |
| 22 | *Personality Disorders/ | 11,489 |
| 23 | *Psychosis/ | 28,409 |
| 24 | *Paranoid Psychosis/ | 1,089 |
| 25 | exp dementia/ | 93,028 |
| 26 | exp Alzheimer's Disease/ | 56,265 |
| 27 | dementia.tw. | 79,166 |
| 28 | alzheimer.tw. | 16,222 |
| 29 | 10 or 11 or 12 or 13 or 14 or 15 or 16 or 17 or 18 or 19 or 20 or 21 or 22 or 23 or 24 or 25 or 26 or 27 or 28 | 1,050,286 |
| 30 | exp Health Care Access/ | 9,861 |
| 31 | (access* or utilis* or utiliz* or uptake* or availab* or service* or pathway* or refer*).tw. | 1,133,781 |
| 32 | engag*.ti. | 27,464 |
| 33 | 30 or 31 or 32 | 1,155,416 |
| 34 | (intervention* or innovation* or improv* or trial*).tw. | 1,083,963 |
| 35 | exp "Systematic Review"/ | 836 |
| 36 | exp Meta Analysis/ | 5,407 |
| 37 | (systematic review* or scoping review* or rapid review* or narrative review* or meta-analysis or evidence synthes#s).ti. | 58,033 |
| 38 | 35 or 36 or 37 | 60,704 |
| 39 | 9 and 29 and 33 and 34 and 38 | 0 |

An additional search focused search was conducted in PsycINFO using the following terms (cultur* or adapt* or modif* or target* or inform* or specific* or tailor* or sensitive).ti and 61 citations were retrieved and screened.

## Footnotes

1 Defined in the Glossary

2 Defined in the Glossary

3 Defined in the Glossary

4 Community coalition-driven interventions involve using “coalitions that include representatives of target populations to plan and implement interventions for community level change.” (Anderson et al. 2015, p. 1)

5 Problem Management Plus is “a transdiagnostic intervention that involves five sessions that teach people skills” (Akhtar et al. 2021, p. 72)

